# Non-Small Cell Lung Cancer Patients Treated with Anti-PD1 Immunotherapy Show Distinct Microbial Signatures and Metabolic Pathways According to Clinical Outcomes

**DOI:** 10.1101/2022.10.05.22280734

**Authors:** David Dora, Balazs Ligeti, Tamas Kovacs, Peter Revisnyei, Gabriella Galffy, Edit Dulka, Dániel Krizsán, Regina Kalcsevszki, Zsolt Megyesfalvi, Balazs Dome, Glenn J. Weiss, Zoltan Lohinai

## Abstract

**Background:** Clinical outcomes of immune checkpoint inhibitor (ICI)-treated non-small cell lung cancer (NSCLC) patients might be associated with the gut metagenome.

**Methods:** Sixty-two Caucasian advanced-stage NSCLC patients treated with anti-PD1 immunotherapy were included. Gut bacterial signatures were evaluated by metagenomic sequencing and correlated with progression-free survival (PFS), PD-L1 expression defined by immunohistochemistry, and other clinicopathological parameters. The predictive role of PFS-related key bacteria was confirmed with multivariate statistical models and validated on an additional patient cohort (n=60). The functional microbial signature of distinct patient groups were determined by analyzing metabolic pathways. A random forest (RF) machine learning algorithm was used to assess the predictive power of taxonomic and metabolic microbial signatures.

**Results:** Alpha-diversity showed no significant difference in any comparison. However, there was a significant difference in beta-diversity between patients with long- (>6 months) vs. short (≤6 months) PFS and between chemotherapy (CHT)-treated vs. CHT-naive cases. Short PFS was associated with increased abundance of Firmicutes (F) and Actinobacteria phyla, whereas elevated abundance of Euryarcheota was specific for low-PD-L1 expression. F/Bacteroides (F/B) ratio was significantly increased in patients with short PFS. Multivariate analysis revealed an association between Alistipes shahii, Alistipes finegoldii, Barnesiella visceriola, and long PFS. In contrast, Streptococcus salivarius, Streptococcus vestibularis, and Bifidobacterium breve were associated with short PFS. Taxonomic profiles performed superiorly in predicting PFS (AUC=0.74), while metabolic pathways including Amino Acid Synthesis and Fermentation were better predictors of PD-L1 expression (AUC=0.87).

**Conclusion:** The specific metagenomic features of the gut microbiome including bacterial taxonomy and metabolic pathways are suggestive of ICI efficacy and PD-L1 expression in NSCLC patients.

## INTRODUCTION

The combination of anti-PD1 immune checkpoint inhibitor- (ICI) and chemotherapy (CHT) represents the backbone for current combination strategies in the front-line setting of advanced-stage non-small cell lung cancer (NSCLC) patients. Nevertheless, even today, only about 20% of all NSCLC patients exhibit stable disease or respond properly to immunotherapy (IT), and only a limited number of patients experience durable benefits [1,2]. In recent years, multiple studies have shown that the gut microbiome might influence ICI efficacy, and CHT might modulate the gut flora. CHT enhances IT efficacy and acts through cancer neoantigen production that T-cells might recognize [3]. Antigens of commensal microbiota can pass through the intestinal barrier and result in T-cell priming, stimulation of cytokine and interferon production, and Toll-like receptor activation through the gut-lung axis [4,5]. This phenomenon is called molecular mimicry, where epitopes produced by microbial species in the gut as part of their natural gene expression programs can resemble tumor neoantigens promoting “autoreactive” T-cells and potent anti-tumor immunity [6].

A key role for Bacteroidales in the immunostimulation associated with ICI was revealed recently [7,8], while others showed a connection between the composition of the gut microbiome and ICI efficacy in malignant melanoma [9,10,11,12], kidney- [13], colorectal and gastrointestinal cancers [14,15], and NSCLC [16,17], or in syngeneic mouse models [18,19]. In a large-scale, multi-cancer cohort of epithelial malignancies, fecal microbiota transplantation (FMT) was also shown to increase ICI efficacy in experimental animals [20].

PD-L1 signaling affects gut mucosa tolerance [21]. However, it is still unclear if there is a direct linkage between the host microbiome and tumor PD-L1 expression. Jang and colleagues hypothesize that metagenomic signatures interact with the tumor and its immune microenvironment, possibly through the microbiome-gut-lung axis [22]. Despite the well-established positive predictive value of tumor PD-L1 immunohistochemistry (IHC) expression in NSCLC [23], clinical evidence shows that more than 50% of PD-L1 high expressing patients might not respond to PD-1/PD-L1 blockade [24]. Altered gut microbiome was also associated with ICI toxicity [25] and other treatments, such as antibiotics [26], steroids [27] or proton-pump inhibitors (PPI) [28]. Others reported that the administration of CHT profoundly disrupts the gut microbiome and affects adverse event frequency, progression-free- (PFS), and overall survival (OS) of cancer patients [29,30].

Given the high variance in response rates concerning immunotherapy, there is an unmet need to discover innovative biomarkers to efficiently select patients for ICI treatments. In our study we analyzed the gut microbiome of 62 immunotherapy-treated NSCLC patients using shotgun metagenomics. In order to reveal predictive bacterial signatures, we have evaluated the bacterial diversity, taxonomy, and metagenome pathways according to PFS, IHC-defined PD-L1 expression, and other clinicopathological parameters such as the front-line CHT-treatment, ICI-related toxicities, and the effects of antibiotic-, antacid- and steroid therapy. To our knowledge, this is the most comprehensive fecal metagenome analysis of Caucasian lung cancer patients treated with anti-PD ICI.

## METHODS

### Study population and treatments

All patients were diagnosed with advanced-stage NSCLC (i.e., stage IIIB/IV). Specifically, we enrolled patients with histologically confirmed adenocarcinoma (ADC), squamous cell carcinoma, and NSCLC not otherwise specified (NSCLC-NOS). The clinical TNM (Tumor, Node, Metastasis) stage was determined according to the Union for International Cancer Control (8th edition) at the time of diagnosis. Clinicopathological data included age at the time of diagnosis, gender, disease stage, smoking pack year (PY), line of immunotherapy (first-line (CHT-naïve) vs. subsequent line (CHT-treated)), tumor PD-L1 expression (IHC, <50% vs. ≥50%), the occurrence of treatment-related adverse events (trAEs, toxicity) and PFS. For trAEs the Common Terminology Criteria for Adverse Events (CTCAE) v5.0 was applied. In line with others [17], ICI-treated patients with complete response (CR), partial response (PR), or stable disease (SD) lasting for at least six months were classified as individuals with long PFS (>6 months) if they achieved. Likewise, patients who experienced progressive disease within six months of treatment initiation were classified as individuals with short PFS (≤6 months). Of note, this classification estimates better the long-term benefits and is more accurate and rigorous in estimating disease progression for a subset of patients who suffer pseudo-progression. PFS was defined as the elapsed time from the commencement of ICI therapy to disease progression according to the aforementioned RECIST 1.0 criteria. The date of last follow-up included in this analysis was 1^st^ of December, 2021. Treatments across all centers were conducted under the current National Comprehensive Cancer Network guidelines. Treatments including antibiotics-, steroids-, and antacids such as proton pump inhibitor and histamine-blocker administration (PPI/H-blocker) were also included. Antibiotic (AB) use, steroid- and PPI/H-blocker treatments prior to the initiation ICI therapy (60 days before) were as well recorded. All patients were assessed with 0-1 ECOG performance status at the initiation of IT.

**STable 1** shows the systemic therapy that patients received at the time of sampling, approved by the Institutional Oncology Teams between 2018 and 2019 at the National Koranyi Institute of Pulmonology, Budapest, Hungary, and at the County Hospital of Pulmonology, Torokbalint, Hungary. **STable 2** shows the comparison of first-line (CHT-naïve) vs subsequent-line (CHT-treated) patients. Patients treated with durvalumab (n=7) and atezolizumab (n=4) participated in phase III clinical trials included in this cohort. The sample collection is not a therapeutic intervention and does not require listing on clinicaltrials.gov.

This study was carried out under the World Medical Association’s Helsinki Declaration study criteria. The national ethics commission officially accepted the study (Hungarian Scientific and Research Ethics Committee of the Medical Research Council (ETTTUKEB-50302-2/2017/EKU)). All patients who participated in/were recruited for the study gave their written consent. After data collection, patient IDs were removed so none of the included individuals can be recognized directly or indirectly.

Processing of fecal samples, DNA extraction, sequencing steps, and the methodology of taxonomic and metagenome pathway assessment, PD-L1 IHC, statistical analyses, and machine learning approach are described in **Supplemental Methods**.

## RESULTS

A total of n=62 consecutive advanced-stage NSCLC patients treated with ICI were enrolled in our study cohort. (**Table 1**). There were 46 patients with long- and 16 with short PFS. 30 patients received first-line IT (CHT-naïve), and 32 patients received ICIs in a subsequent-line (CHT-treated) (**STable 2**). We also analyzed an ICI-treated Validation cohort of advanced-stage NSCLC patients treated between 2017 and 2018 at the same institutions with the same treatments and guidelines to validate the relative abundance of key bacterial species associated with long- or short PFS (n=60, **STable 3**). **STable 4** shows the comparison in clinical parameters between the Discovery and Validation cohorts, where the proportion of PD-L1 low IHC expressing patients (p=0.001) and CHT-treated patients (p<0.001) were significantly higher compared to the Discovery cohort. **SFig 1A** shows the study design on a flow chart. In the Discovery cohort, CHT-naive and PD-L1-high patients showed significantly better PFS compared to CHT-treated (p=0.0016) and PD-L1-low patients (p=0.0041), respectively (**SFig 1C-D**). No significant difference was detected in PFS according gender (p=0.055, **SFig 1B**), toxicity (p=0.872, **SFig 1E**), antibiotic treatment (p=0.247, **SFig 1F**), antacid medication (p=0.88, **SFig 1G**) and steroid treatment (p=0.0894, **SFig 1H**). **STable 5a-b** shows uni- and multivariate Cox hazard regression with the baseline clinical parameters, where PD-L1 status (high vs. low) was the only significant predictor in the multivariate analysis (p=0.005, HR: [3.861]).

**Table 1.** Clinicopathological characteristics of Discovery cohort. *P-values are indicated for Fischer’s exact test, Mann-Whitney test or Welch’s test in case of normality*.

|  | Long PFS<br>N=46 (74%) | Short PFS<br>N=16 (26%) | p-value |
| --- | --- | --- | --- |
| Age (mean) | 59.53 | 61.11 | 0.099 |
| Gender (male) | 26.09% (12) | 50% (8) | 0.119 |
| ICI Response (R) at month 3 | 100% (45) | 31.25% (5) | <0.001 *** |
| PFS (months, median) | 20.83 | 3.48 | <0.001 *** |
| Chemotherapy (treated) | 45.65% (21) | 68.75% (11) | 0.150 |
| PD-L1 IHC (>50%) | 50% (23) | 25% (4) | 0.190 |
| Smoking (pack-years) | 42.87 | 31.87 | 0.166 |
| ICI Toxicity (present) | 30.43% (14) | 21.42% (3) | 0.736 |
| Antibiotics (pre-ICI) | 19.56% (9) | 25% (4) | 0.725 |
| Steroids (pre-ICI) | 34.78% (16) | 46.66% (7) | 0.541 |
| Antacids (pre-ICI) | 58.69% (27) | 60% (9) | >0.99 |

### The composition of bacterial communities differs significantly according to progression-free survival and chemotherapy treatment

First, alpha-diversity was assessed in fecal samples according to PFS, PD-L1 expression, and CHT treatment for all major phylogenetic levels, including phylum, class, genus, and species. Shannon index showed no significant differences in any comparisons (**Fig 1A-C**). UMAP plot and beta-diversity analyses showed that short PFS patients represent a significantly different bacterial composition compared to long PFS patients (p<0.001, **Fig 1D**), whereas patient groups do not differ significantly according to PD-L1 expression (p=0.508, **Fig 1E**). Interestingly, front-line CHT-treated patients exhibit a significantly different bacterial community than CHT-naive patients (p=0.015, **Fig 1F**).

**Figure 1.**
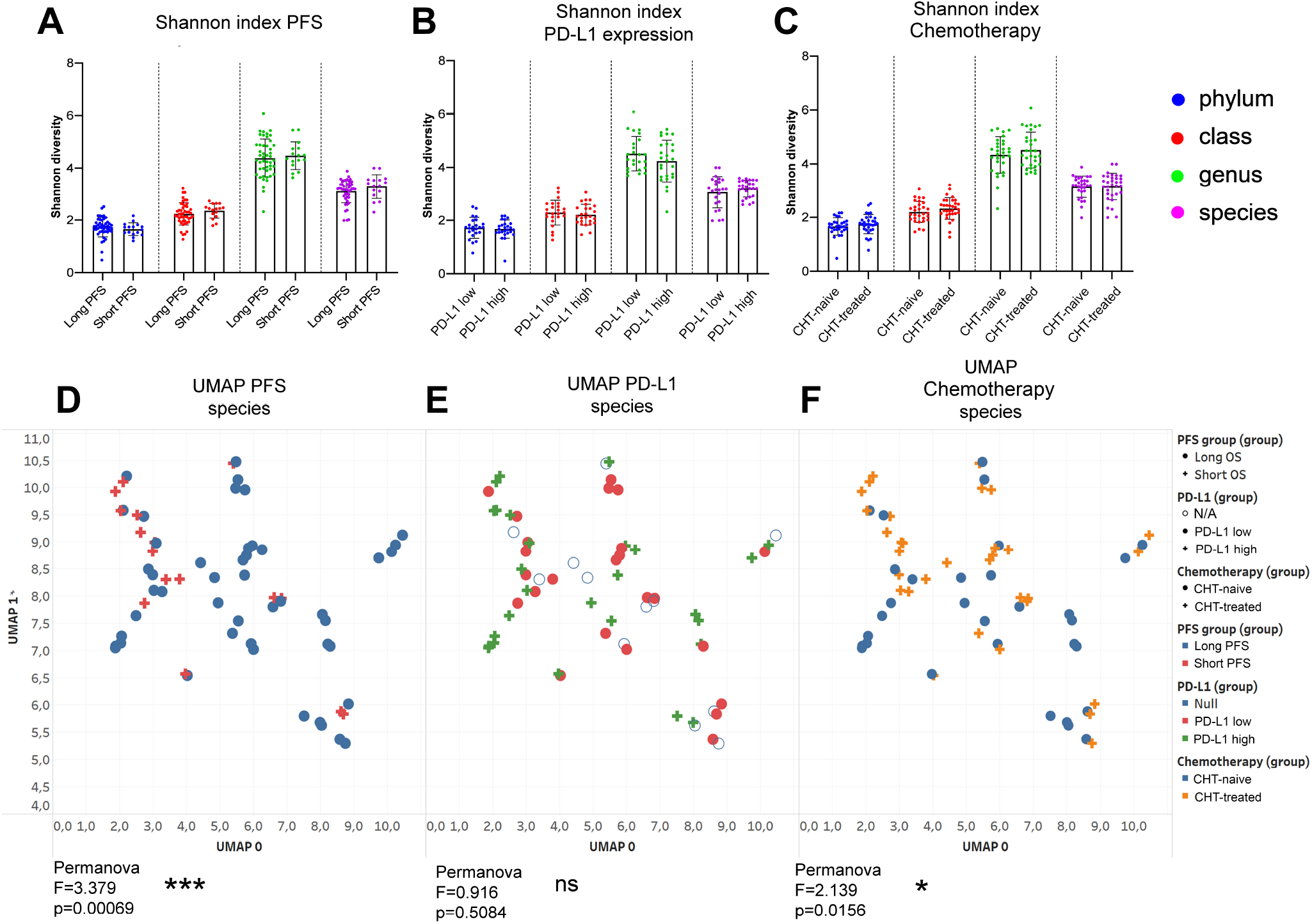
Alpha diversity and Composition of bacterial communities (beta diversity) Shannon diversity index was calculated at phylum, class, genus and species taxonomy level according to long vs short PFS (cut-off 6 months), PD-L1 expression (≥50% high, <50% low) and front-line chemotherapy (CHT)-treatment prior to ICI. There was no significant difference in Shannon diversity index regarding PFS (**A**), PD-L1 expression (**B**) and CHT-treatment (**C**) using either taxonomic level. Diversity (Shannon and Simpson) indices and p-values for all alpha-diversity comparisons are listed in **STable 3**. Ordination plot using UMAP was generated from normalized, CLR-transformed bacterial abundances in the same comparisons. Permanova analysis was used to assess significant differences between the composition of bacterial communities. There was a significant difference between long- vs short PFS patients (F=3.379, p=0.0006, **D**) and between CHT-treated vs CHT-naive patients (F=2.139, p=0.0156, **F**), whereas no significant difference was detected between PD-L1 high vs PD-L1 low patients (F=0.916, p=0.0156, **D**) regarding the composition of bacterial species. *Metric data are shown as mean and corresponding standard deviation (SD). Statistical significance *P < 0*.*05; **P < 0*.*01, ***P<0*.*001. N/A: data not available*.

### Firmicutes and Actinobacteria phyla are more abundant in patients with short PFS, along with an increased F/B ratio

**Figure 2A** shows phylum composition for every patient, where all phyla with CLR≥-1 were included. For further analysis, we removed all non-bacterial taxonomic units such as fungi (Ascomycota), protozoa (Apicomplexa), viruses, Chordata (human host), and plants (Streptophyta). Firmicutes, Bacteriodetes, Proteobacteria, Actinobacteria, and Verrucomicrobia represent high abundance phyla (CLR≥0), whereas low abundance phyla (0>CLR≥-1) included Euryarcheota, Cyanobacteria, Spirochetes, and Tenericutes in the gut microbiome of our patient cohort (**Fig 2B**). Firmicutes (p=0.011, **Fig 2C**) and Actinobacteria (p=0.004, **Fig 2D**) were significantly more abundant in patients with short PFS. Euryarcheota were significantly more abundant in PD-L1-low patients than in PD-L1 high expressors (p=0.001, **SFig 2A**) and Cyanobacteria were significantly more abundant in CHT-naive patients (compared to CHT-treated, p=0.029, **SFig 2B**). An increased Firmicutes/Bacteroidetes (F/B) ratio was associated with short-term PFS (p=0.013, **Fig 2E**). Furthermore, we detected an increased Actinobacteria to Proteobacteria ratio (A/P ratio) in patients with short PFS (p=0.011, **Fig 2F**).

**Figure 2.**
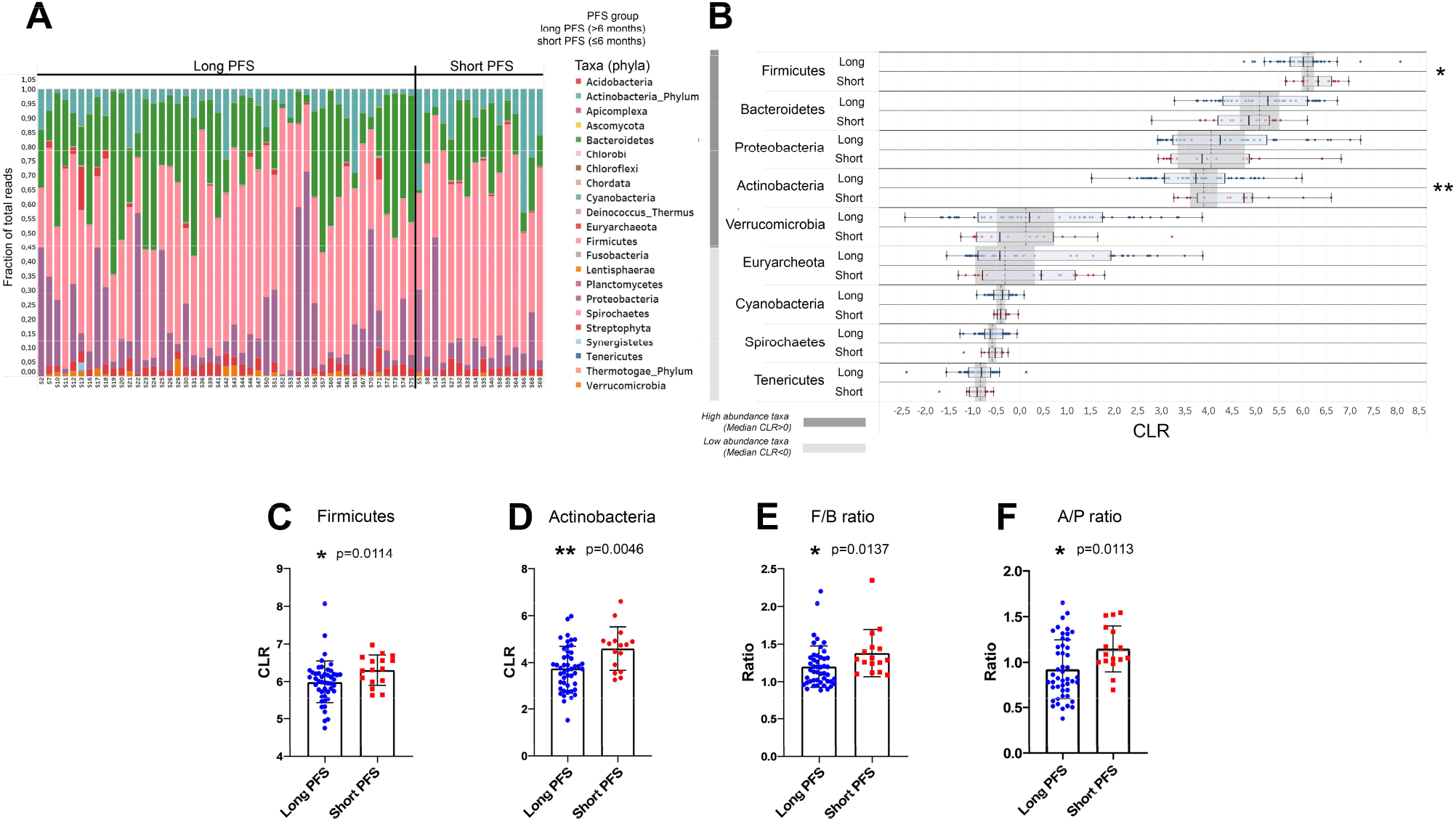
Differentially abundant Phyla and outcomes. Phylum composition (fraction of total reads) of all patients in the Discovery cohort value is shown in the stacked bar chart (**A**). After removing phyla with minimal abundance (CLR<-1) and excluding non-Bacteria and non-Archea taxa, Wilcoxon rank-sum tests were performed to compare their abundance in patients with long- vs short PFS (**B**). Firmicutes and Actinobacteria are significantly more abundant in patients with short PFS vs long PFS (4.704 vs 3.612 and 6.213 vs 5.935, p=0.0114 and p=0.0046, respectively, **C and D**). Moreover, both Firmicutes/Bacteroidetes (F/B, 1.294 vs 1.132, p=0.0137, **E**) and Actinobacteria/Proteobacteria ratio (A/P, 1.068 vs 0.827, p=0.0113, **F**) were significantly increased in patients with short PFS. Regression curves show significant negative correlation between PFS (months) and abundance (CLR) of phyla Actinobacteria (r=−0.35, p=0.004, **G**) and Firmicutes (r=−0.32, p=0.011, **H**). *Metric data are shown as mean and corresponding standard deviation (SD). Statistical significance *P < 0*.*05; **P < 0*.*01, ***P<0*.*001*.

### Multiple genera and species are differentially abundant in patients with long- vs. short progression-free survival

We performed Wilcoxon rank-sum test for all detected genera and species (high abundance: CLR≥0; low abundance 0>CLR≥-1) 11 genera and 26 species showed a significant difference in abundance according to PFS (**Fig 3A, B**). We labeled these taxa as key genera and species. High-abundance genera Streptococcus (p=0.017), Bifidobacterium (p=0.002), Colinsella (p=0.024) and Lactobacillus (p=0.043) were more abundant in patients with short PFS, whereas Alistipes (p=0.018), Paraprevotella (p=0.031) and Barnesiella (p=0.001) were more abundant in patients with long PFS. Among low-abundance genera, Spiroplasma (p=0.034), Helicobacter (p=0.033), and Buchnera (p=0.021) were rather associated with long-term PFS, while Methanospaera (p=0.02) was more associated with short PFS (**Fig 3A**). Multiple species showed increased abundance in patients with long PFS (**Fig 3B**), from which the most significant taxa included Alistipes shahii (p<0.001), Barnesiella visceriola (p<0.001), Butyricimonas faecalis (p=0.001), Bacteroides sp. A1C1 (p=0.003) and Alistipes finegoldii (p=0.004). Patients with short PFS patients were associated with a significantly increased abundance of variety of Streptococcus species, such as S. salivarius (p=0.002), S. vestibularis (p=0.005), and S. parasanguinis (p=0.01); Bifidobacteria, such as B. longum (p=0.017), B. adolescentis (p=0.006) and B. breve (p=0.007). Moreover, there was an increased abundance of Colinsella aerofaciens (p=0.004) and low-abundance Streptococci in Short PFS patients (**Fig 3B**).

**Figure 3.**
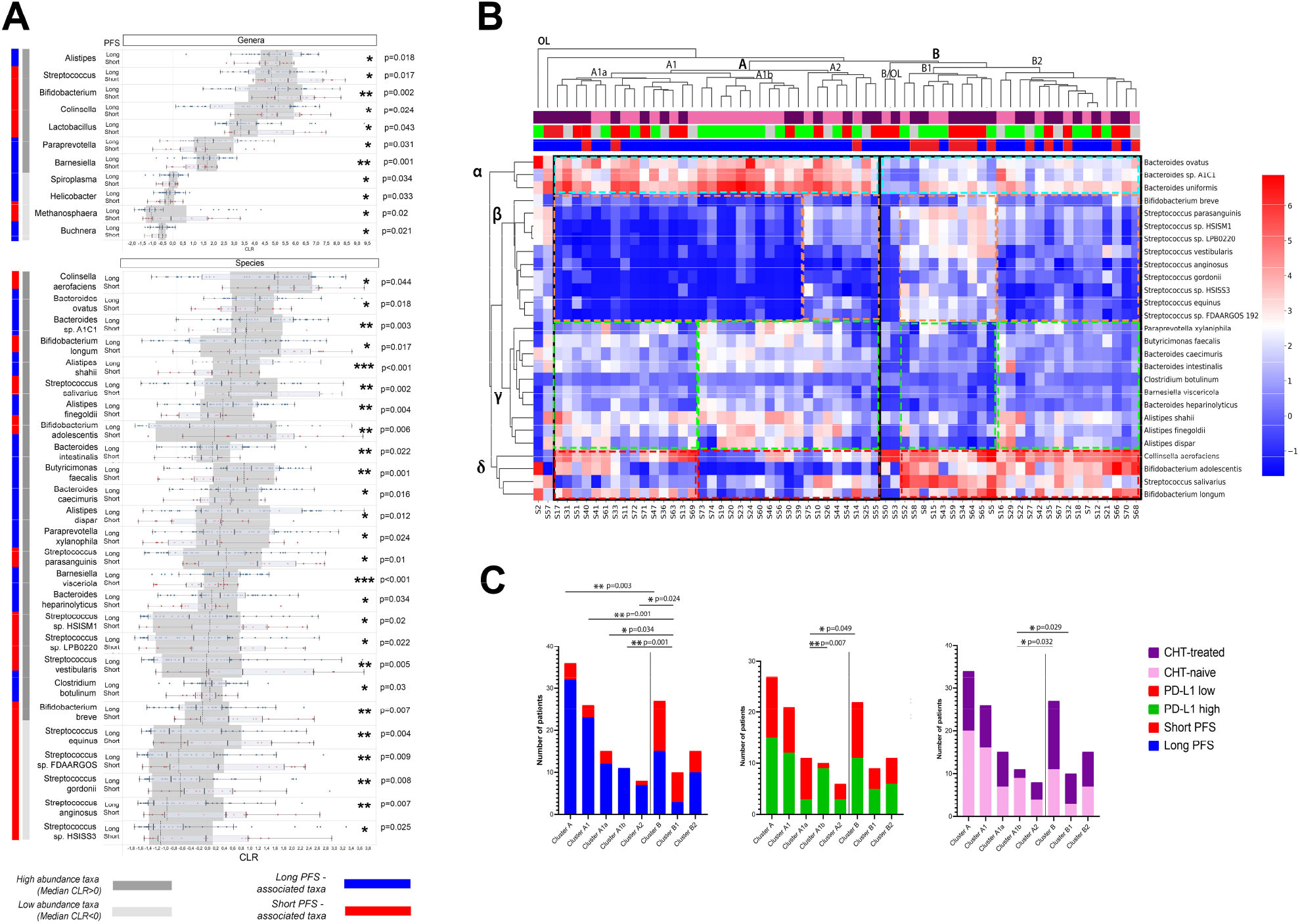
Differentially abundant genera and species and outcomes. Cluster analyses. Horizontal bar charts show the abundance of key genera and species with significantly different abundance between patients with long- vs short PFS, Wilcoxon ranksum test, p<0.05 (**A**). Only genera and species (CLR>-1) from the Bacteria and Archea domains were included in the analyses. High abundance taxa (CLR ≥ 0) and low abundance taxa (0 > CLR ≥ −1) are separately labeled. Heatmap displays Z-scores (blue=low, red=high) for every cell generated from individual CLR-transformed abundances from key species for all patients (**B**). Axis X shows patients (IDs) in the Discovery cohort, whereas indicator bars on top reflect their PFS (red/blue, short vs long), PD-L1 (green/red, high vs low) and front-line CHT-treatment (purple/pink, CHT-treated vs CHT-naive) as previously described (**B**). Axis Y shows key bacterial species clustered to representative groups (**STable 5**). There were two outlier patients (S2, S57), in the whole cohort and two outlier patients in cluster B (S50, S53), who cannot be clustered to any groups. Patient clusters are compared shown in stacked bar charts (**C**) according to their composition of long vs short PFS, PD-L1 high vs low and CHT-treated vs naive patients. Cluster A represents significantly more patients with Long PFS (compared to cluster B, p=0.003, **C**) with an increased abundance of beneficial α and γ bacteria, a decreased abundance of β and a decreased- or variable abundance of δ bacteria; cluster B is characterized by a decreased abundance of α and γ, an increased abundance of δ and an increased or variable abundance of β bacteria. Cluster A1a and A1b represent a PD-L1-low and high subcluster in cluster A, with no significant difference in patients according to PFS. Cluster A1b consists of significantly more CHT-naive patients than cluster B in general. Fisher’s exact test was used to calculate differences among all clusters and subclusters. *Metric data are shown as mean and corresponding standard deviation (SD). Statistical significance *P < 0*.*05; **P < 0*.*01, ***P<0*.*001*.

Significant taxonomical associations with PD-L1 expression and front-line CHT-treatment were revealed using the previous methodology, where multiple genera and species showed a significant difference in PD-L1 expression (**SFig 3A**), including Methanobrevibacter (p=0.002) and Microbacter (p=0.009) or Methanobrevibacter smithii (p=0.013). **SFig 3B** show Receiver Operator Characteristic (ROC) curves and a table for genera with the highest AUC to predict PD-L1 high vs. low expression in patients. M. smithii showed the best AUC to predict PD-L1 phenotype among species (**SFig 3B**). Differentially abundant genera and species in CHT-treated vs. CHT-naive patients are shown in **SFig 3C** and **D**.

CHT-treated patients exhibit a significantly different taxonomic composition from CHT-naive (**Figure 1F**) patients, therefore we evaluated differential abundance between patients with short vs. long PFS in a selected population of CHT-treated patients (**STable 6**). A. shahii, S. salivarius, S. vestibularis, and B. adolescentis still showed differential abundance between patients with long- vs. short PFS. Interestingly, in this selected CHT-treated population, E. coli showed significantly increased abundance in patients with long PFS and R. bicirculans in patients with short PFS but not in the whole cohort.

Next, a heatmap was generated to reveal clusters formed by key bacterial species and patients. Bacteria were clustered into four groups labeled with Greek letters (**Figure 3B**). Cluster α constitutes high-abundance Bacteroides, including B. uniformis, B. sp. A1C1, and B. ovatus. Cluster β includes B. breve and a variety of Streptococci. Cluster γ represents a heterogeneous microbial community with bacteria beneficial for PFS, including Bacteroides, Alistipes, and Butyricimonas species. Short PFS-associated species B. adolescentis, B. longum, C. aerofaciens, and S. salivarius comprise Cluster δ. Bacterial clusters and their representation among patients are described in **STable 7**. Of note, we observed that at least two bacterial clusters are altered in patients with short PFS, compared to patients with long PFS, including underrepresentation of beneficial (cluster α, γ), or overrepresentation of detrimental bacteria (clusters β, δ). Underrepresentation of a singular beneficial- or overrepresentation of a singular detrimental bacterial cluster was not essentially associated with short PFS. Characteristics and statistical comparison of sub-clusters derived from patients relative to PFS, PD-L1 expression, and CHT are shown in **Fig 3C**. Altogether, patients with long PFS are significantly overrepresented in cluster A, and patients with short PFS patients in cluster B (**Fig 3C**).

### Networks of bacterial communities are significantly different in patients with long vs. short PFS

Interaction networks were generated using Spearman’s coefficients among bacterial taxa, including all samples (n=62), and only samples of patients with long (n=46) - or short-PFS (n=16). **Fig 4A** shows legends for network diagrams. In the whole cohort, Actinobacteria vs. Firmicutes showed a moderate positive correlation (r=0.406), whereas there was a moderate negative correlation between Euryarchaeota vs. Firmicutes (r=−0.522), Euryarchaeota vs. Bacteroidetes (r=−0.412) and Bacteroidetes vs. Actinobacteria (r=−0.409) phyla. Proteobacteria and Verrucomicrobia exhibited no significant associations with other phyla (**Fig 4B**). We also compared bacterial correlations in patients with short vs. long PFS. According to differential analysis, where the long PFS group was set as reference, Verrucomicrobia’s correlation has decreased with Firmicutes and increased with Bacteroidetes. Bacteroidetes’ correlation has decreased-, Euryarcheota’s has increased with Actinobacteria (**Fig 4C**). **Figures 4D and E** show a network of key genera in the whole cohort (**D**) and on the Tiffany diagram (**E**). Interestingly, Actinobacteria, such as Bifidobacterium, Actinomyces, and Colinsella, correlate positively with Firmicutes, such as Lactobacillus and Streptococcus. In contrast, Bacteroidetes Butyricimonas, Barnesiella, and Alistipes negatively correlate with key Firmicutes and Actinobacteria genera (**Fig 4D**). The connectivity network and differential network for all genera (CLR>0) are shown in **SFig 4A**-**B**.

**Figure 4.**
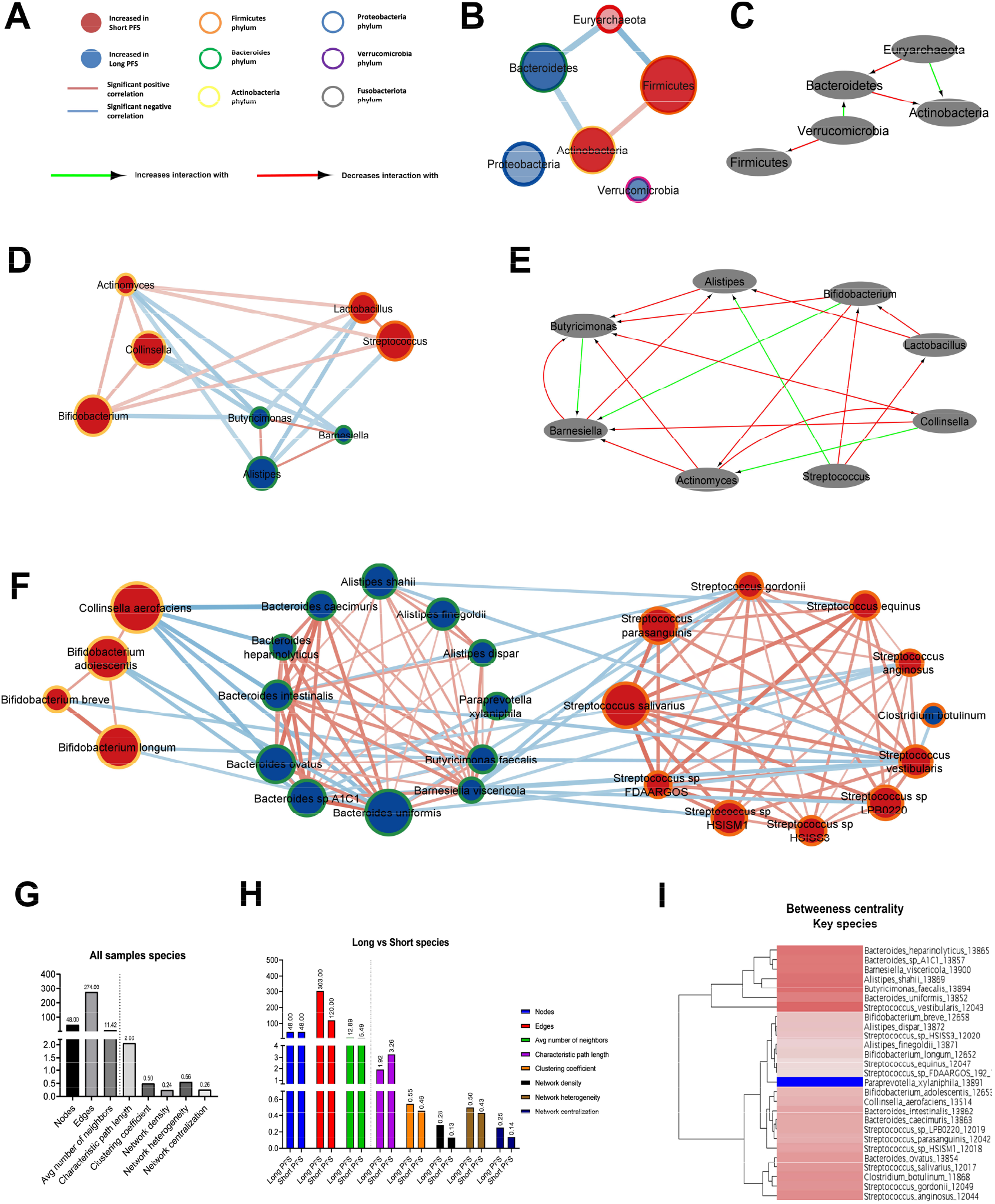
Bacterial networks in patients with long- vs short PFS. Network maps show the correlation, phylogenetic origin and representation in patients with long- vs short PFS of key genera and species. Nodes represent taxa, where association with long (blue)- or short (red) PFS and phylogenetic classification are color-coded (**A**). Size of nodes reflects median CLR value of the taxon in the whole cohort. Edges represent significant (p<0.01) positive (red) or negative (blue) correlations (r > [0.4]) among nodes. Thickness of edges reflect rho values according to Spearman’s correlation coefficients. Correlations r < [0.4] and p ≥ 0.01 are not shown in networks. Nodes and taxa without minimum one significant correlation (r > [0.4], p<0.01) and a minimum median CLR of 0 (genera) or −1 (species) are not shown in networks (**B**,**D**,**F**,**G**). Using the Diffany network analyzer module from Cytoscape, differential analysis of long- vs short PFS networks were performed, where red arrows depict a decreasing level of association-, while green arrows denote an increasing level of association between two taxa in bacterial networks. Diffany diagrams use the long PFS population as reference point and shows how the short PFS population differs from it (**C**,**E**,**H**). Long PFS patient-networks were used as reference for Diffany graphs, so diagrams depict how the network differs in patients with short PFS. Panels show whole cohort network and Diffany diagram for phyla (**B**,**C**), and for key genera (**D**,**E**). Panel **F** shows whole cohort network for key species. Number of nodes, edges, average number of neighbors, characteristic path length between nodes, clustering coefficient, density, heterogeneity and centralization of networks are shown in the whole cohort (**G**) and compared between long- and short PFS patient networks (**H**). Dendrogram depicts clusters in the whole cohort species, revealing a cluster (A) with high betweenness centrality and a cluster (B) with relatively low betweenes centrality (**I**).

**Figure 4F** depicts the whole cohort network using key PFS-associated species, where Bacteroidetes species strongly correlate with each other and show a strong negative correlation with C. aerofaciens. Streptococcus species showed a similar reciprocal correlation within their taxonomic niche, whereas S. vestibularis, S. anginosus, and S. gordonii were negatively correlated with multiple Bacteroidetes species (**Fig 4F**). **SFig 4C** shows the differences between networks generated from patients with short- and long PFS.

The whole cohort network’s characteristics and comparison of key species’ long- vs. short PFS networks are shown in **Fig 4G-H**. The Long PFS network is more interconnected with a higher number of edges for the same number of nodes, decreased characteristic path length, increased network density, heterogeneity, and centralization (**Fig 4H**). Dendrogram displays nodes (taxa) according to betweenness centrality. Cluster A bacteria, such as S. vestibularis, B. uniformis, B. faecalis, A. shahii, B. visceriola, B. sp. A1C1, and B. heparinolyticus are considered central hubs in the PFS-related gut microbiome with a multitude of connections. Paraprevotella xylanophila is the most isolated species, with the lowest number of associations with other species from cluster B (**SFig 4I**).

### Taxa associated with ICI toxicity and history of Antibiotic-, Steroid, and PPI/H-blocker treatments

Next, we aimed to reveal differentially abundant taxa according to the presence of ICI adverse events (toxicity) and medications taken before IT, including antibiotics, antacids, and steroids. Treatment toxicity was associated with a slight decrease in the abundance of genera Absiella and Blautia (**Fig 5A**) and a pronounced increase in the abundance of Prevotella dentalis (**Fig 5A’**). Antibiotic treatment affected multiple taxa, significantly decreasing the abundance of Anaerostipes, Christensenella, Longibaculum, Lachnospira, Anaerostipes Hadrus, and Erysipelothrichaceae bacterium GAM147 (**Fig 5B-B’**). In contrast, Eggerthella (E) and E. lenta, Bifidobacterium bifidum, and Bacteroides xylanisolvens were significantly overrepresented in patients with a history of antibiotic treatment (**Fig 5B-B’**). Antacid medication was associated with a modest decrease in Synergistetes phylum (**Fig 5C**) and a marked increase in species Streptococcus (S) equinus, S. parasanguinis, and S. salivarius (**Fig 5C’**), whose increased abundance was also detected in short PFS patients (**Fig 3A**). The phylum Proteobacteria and its prominent genus Eschericia (E) and species E.coli were significantly more abundant in steroid-treated patients, similarly to Longibaculum and Erysipelothrichaceae bacterium GAM147 (**Fig 5D-D’**).

**Figure 5.**
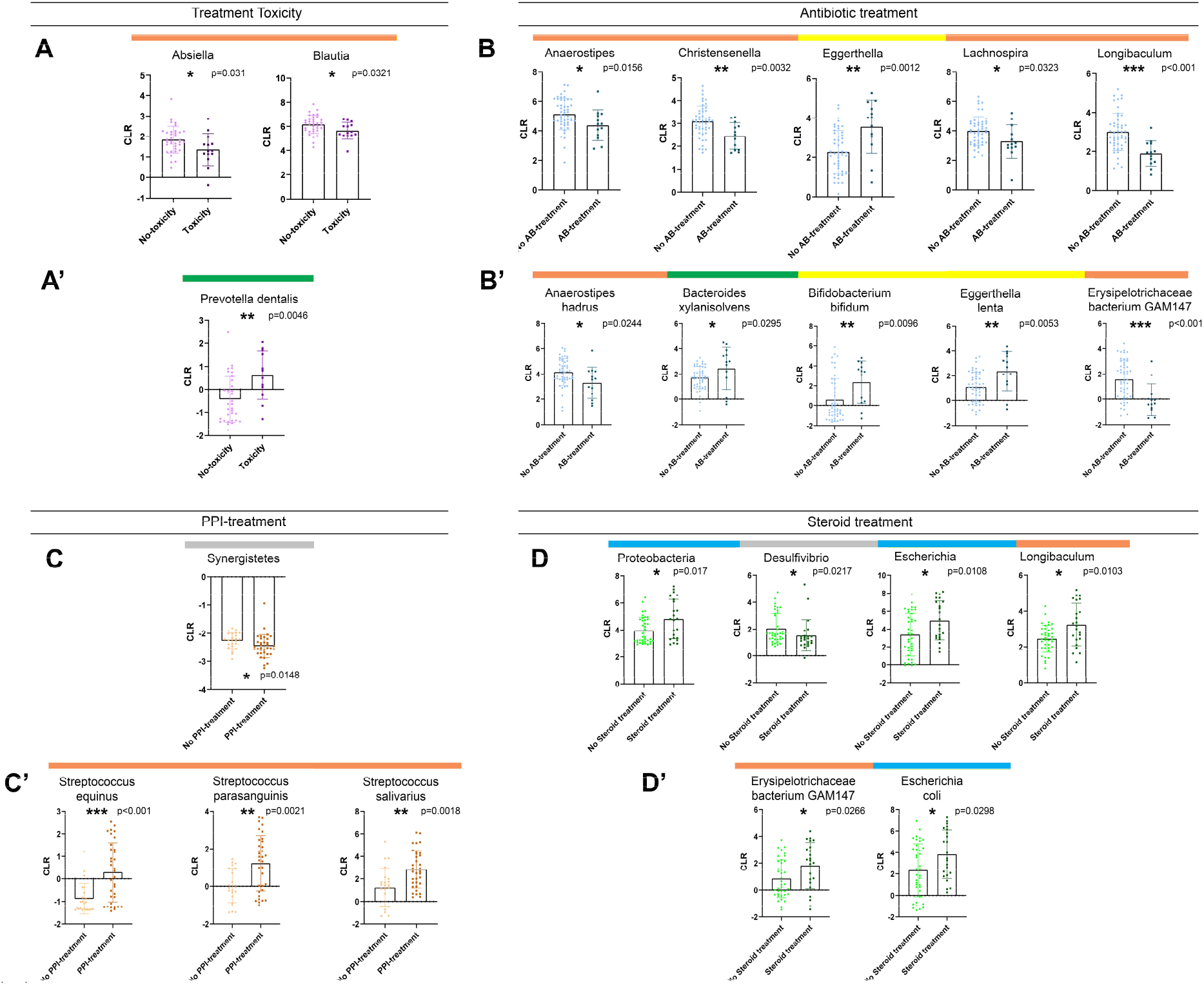
Differentially abundant taxa according to ICI-toxicity and Antibiotic-, Antacid-, or Steroid treatment before Immunotherapy. Bar charts show the CLR-normalized abundance of taxa with significantly different abundance (Wilcoxon ranksum test, p<0.05) according to ICI-toxicity (**A-A’**), Antibiotic (AB)-treatment prior IT (**B-B’**), Antacid treatment prior or during IT (**C-C’**) and steroid treatment prior or during IT (**D-D’**). Only genera and species (CLR>-1) from the Bacteria and Archea domains were included in the analyses. Genera Absiella and Blautia showed significantly decreased abundance in patients with no ICI-toxicity (p=0.031 and p=0.032, **A**), whereas species Prevotella dentalis was significantly more abundant in patients with ICI-toxicity (p=0.0046, **A’**). Genera Anaerostipes (p=0.0156), Christensenella (p=0.0032), Lachnospira (p=0.0323) and Longibaculum (p<0.001) showed significant depletion, whereas Eggerthela (p=0.0012) was significantly increased in AB-treated patients (**B**). Regarding species, Anaerostipes hadrus (p=0.0244) and Erysipelothrichaceae bacterium GAM147 (p<0.001) showed a significantly decreased abundance, whereas Bacteroides xylanisolvens (p=0.0295), Bifidobacterium Bifidum (p=0.0096) and Eggerthella lentha (p=0.0053) showed a significantly increased abundance in AB-treated patients (**B’**). Low-abundance phylum Synergistetes showed a slight, but significant decrease in patients with a history of- or ongoing Antacid medication (**C**), whereas Streptococci (S) S. equinus (p<0.001), S. parasanguinis (p=0.0021) and S. salivarius (p=0.0018) exhibited significantly increased abundances in Antacid treated patients (**C’**). Steroid treatment prior or during IT was associated with a significantly increased abundance of Proteobacteria phylum (p=0.017) and genera Escherichia (p=0.0108) and Longibaculum (p=0.0103), whereas abundance of genus Desulfovibrio (p=0.0217) was significantly decreased in the same group of patients (**D**). Two species showed significantly increased abundance in steroid-treated patients: Erysipelothrichaceae bacterium GAM147 (p=0.0266) and Escherichia coli (p=0.0298) (**D’**). *Horizontal colored bars above charts show the phylogenetic origin of taxa (see description in Figure 4A)*.

### Multivariate statistical models

ROC curves were generated, and AUC values were measured to reveal the predictive power of key bacterial taxa regarding long or short PFS and to assess their prospect as clinical tests in predicting ICI efficacy (**Figure 6A**). Results of Wilcoxon tests ROC-analyses are shown in **STable 8**. B. faecalis, S. parasanguinis, B. breve, and B. visceriola represent high-specificity, but mediocre sensitivity taxa, while S. vestibularis, S. salivarius, A. finegoldii, B. adolescentis, and B. sp. A1C1 represent a high-sensitivity, but mediocre specificity taxa. A. shahii is the only species showing strong specificity and sensitivity to predict PFS.

**Figure 6.**
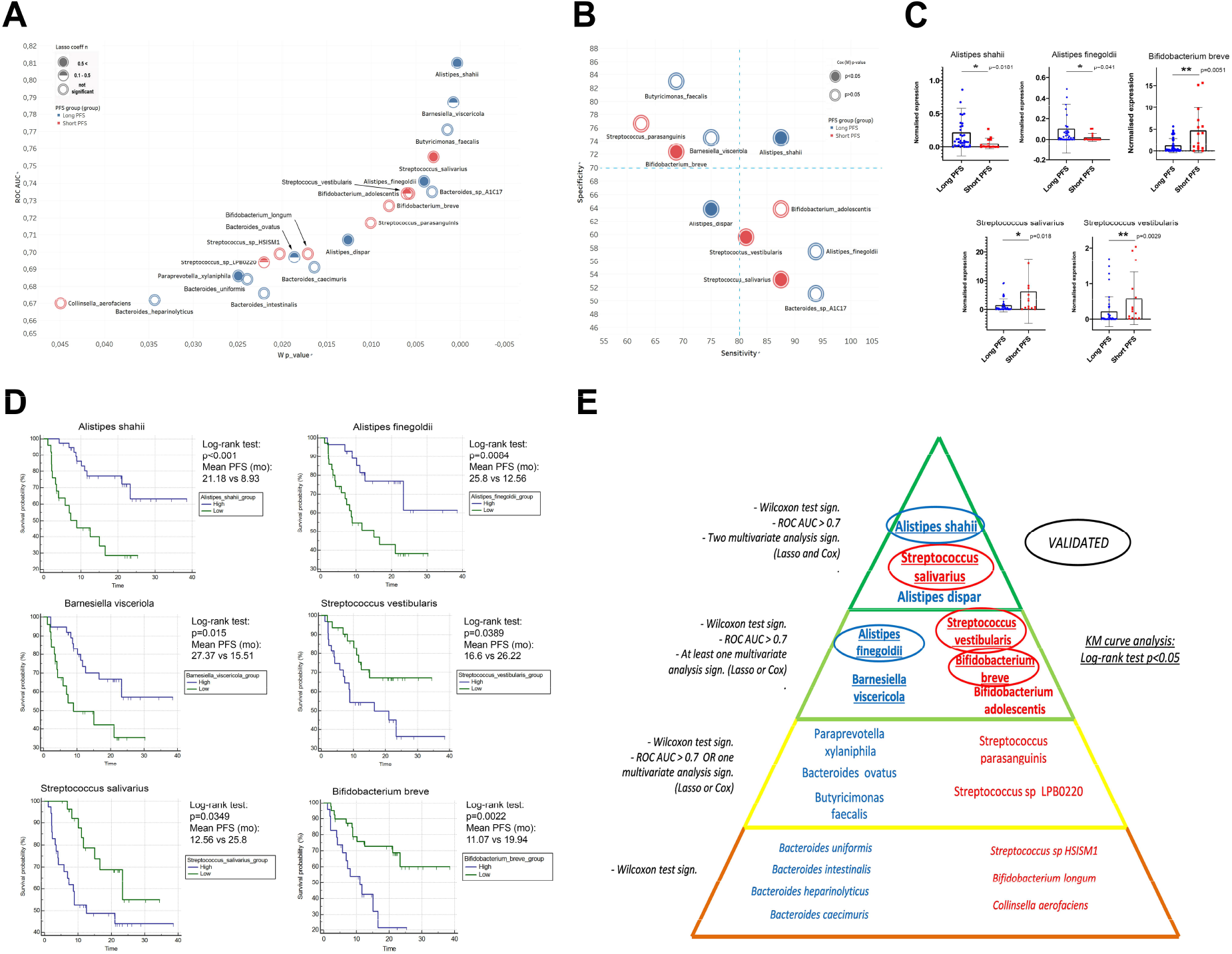
Multivariate and survival analysis. XY chart visualize key species according to AUC value from ROC analysis (axis Y) and p-value of Wilcoxon rank sum test (axis X) relative to the binary classification of long vs short PFS (**A**). Association of bacterial species with PFS is depicted in red (short) and blue (long). The normal value of Lasso coefficient is depicted in full (>0.5), half (0.1 - 0.5) and empty (not significant) spheres (**A**). Sensitivity-Specificity chart plots the same species, where specificity (axis Y), and sensitivity (axis X) are denoted in percentage (**B**). Association of bacterial species with PFS is depicted in red (short) and blue (long). The p-value from Cox (M) analysis is depicted in full (p < 0.05) and empty (p ≥ 0.05) spheres (**B**). Abundance of A. shahii (p=0.0181) and A. finegoldii (p=0.041) was significantly increased in patients with long- (vs short) PFS, whereas abundance of S. salivarius (p=0.018), S. vestibularis (p=0.0029) and B. breve (p=0.0051) was significantly increased in patients with short- (vs long) PFS according to validation on an additional patient cohort. There was no significant difference in the case of B. adolescentis (p=0.692). There were no abundance data available for B. visceriola, and A. dispar in the validation cohort’s Metaphlan2 dataset (**C**). Hierarchical pyramid-model shows key bacterial species according to long (blue) vs short (red) PFS (**E**): Level 1: Significant (p<0.05) Wilcoxon Rank sum test; Level 2: Level 1 requirement AND at least fair (AUC>0.7) ROC curve OR at least one multivariate analysis (Lasso or Cox) with significant result; Level 3: Level 1 requirement AND at least fair (AUC>0.7) ROC curve AND at least one multivariate analysis (Lasso or Cox) showing significant result; Level 4: Level 1 requirement AND at least fair (AUC>0.7) ROC curve AND both multivariate analyses (Lasso and Cox) showing significant results (**E**). Bacteria on Level 3 and 4 were validated on an additional patient cohort (**C**) and KM curves were generated with cut-offs derived from ROC analysis for all species on levels 3 and 4 (**D**). A. shahii, A. finegoldii and B. visceriola - high patients showed significantly increased PFS compared to patients with low abundance of these species. Patients with increased abundances of S. vestibularis, S. salivarius and B. breve exhibited decreased PFS, compared to patients with low abundance of these species. Panel **D** displays KM curves with median PFS in months, p-values of Log-rank tests and patient numbers at risk in every comparison. *Encircled: Successfully validated; Underlined: KM curve analysis shows significant difference (Log-rank test)*

Next, we established a gut bacteria-based multivariate model for predicting IT response, where Lasso regression and Cox proportional hazard regression were used. Univariate- (U) and multivariate (M) models were used for Cox regression. Univariate analysis (Cox (U)) was performed for all key species (**Table 2**) with a significant, ROC curve (AUC>0.7). In multivariate Cox models, key species with significant (p<0.05) Cox (U) predictive values and confounding parameters such as gender, PD-L1 expression, and CHT-treatment were included (**Fig 6A-B**). To exemplify the hierarchy of key species according to their association with PFS, a pyramid model is shown in **Fig 6E**.

**Table 2.**
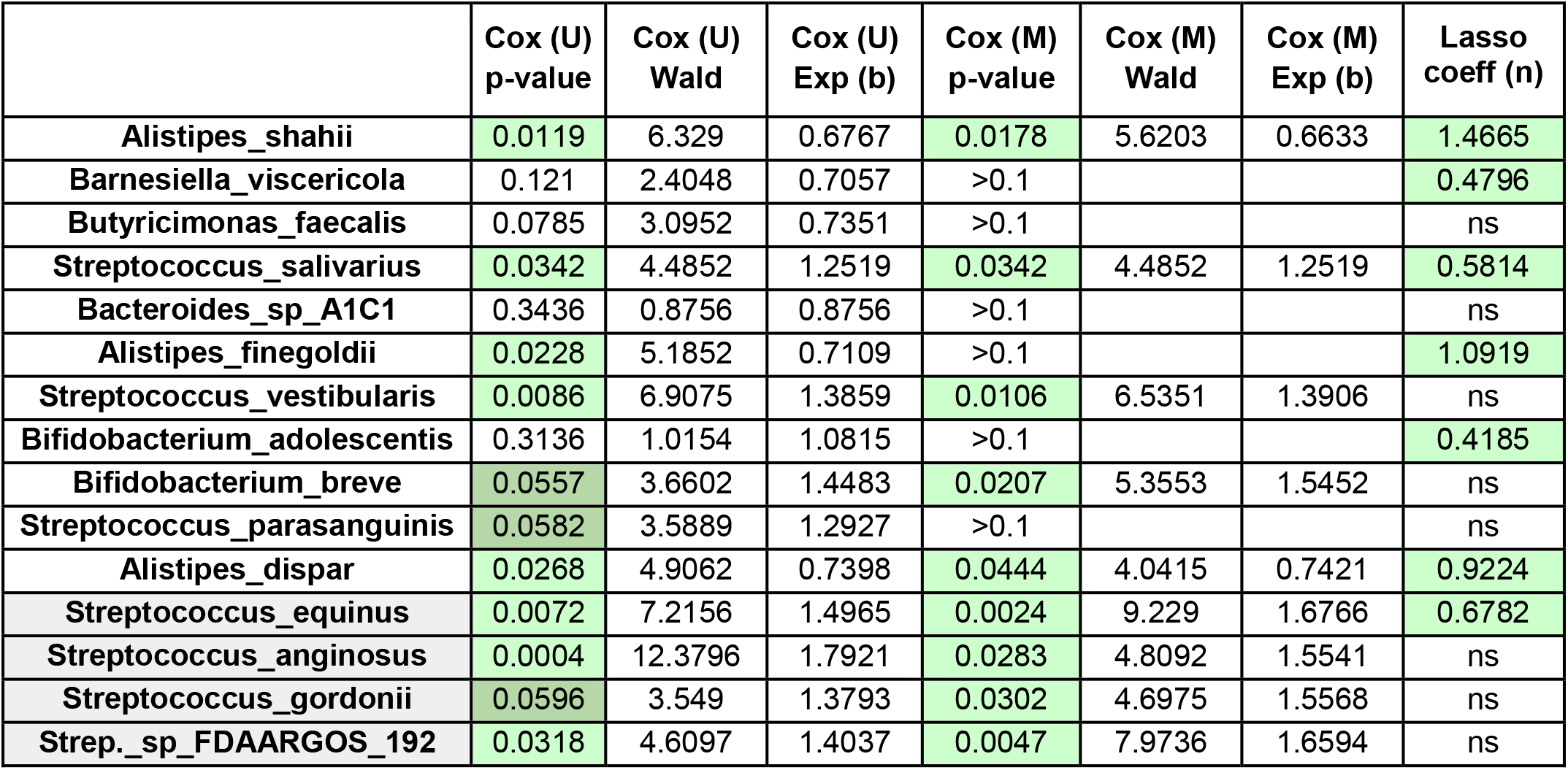
Results of Uni- and Multivariate Cox regression for key species. Green color represents significant (<0.05) p-values and dark green color shows only a trend. Cox (M) analysis was performed only for taxa with a significant result (p<0.1) in Cox (U) analysis. Grey colored taxa are low-abundance. For Lasso regression, Lasso coeff (n) is only shown for significant (p<0.05) predictors

An additional patient cohort (n=60, **STable 3**) was used for validation. According to our hierarchical pyramid model, we aimed to validate level 3 and 4 species on our validation cohort, where at least one multivariate regression model (Lasso or multivariate Cox) showed a significant predictive role. We confirmed species A. shahii (p=0.0181) and A. finegoldii (p=0.041) that were significantly increased in patients with long- (vs. short) PFS, and species S. salivarius (p=0.018), S. vestibularis (p=0.0029) and B. breve (p=0.0051), that were significantly increased in patients with short- (vs. long) PFS (**Fig 6C, E**). Abundance of B. adolescentis did not show a significant difference in the validation cohort (**SFig 6A**). Cut-off values were generated based on the ROC curves, and KM analysis was performed for level 3 and 4 species. An increased abundance of A. shahii (p<0.001), A. finegoldii (p=0.0084), and B. visceriola (p=0.015) was associated with significantly improved PFS. Conversely, increased abundance of S. salivarius (p=0.0389), S. vestibularis (p=0.0349), and B. breve (p=0.0022) was associated with significantly shortened PFS (**Fig 6D, E**). According to KM analysis, B. adolescentis and A. dispar showed no significant difference in PFS (**SFig 6B-C**). Results of Cox regression for key phyla and genera are shown in **STable 9**. Lasso regression underpinned the negative predictive role of both Firmicutes and Actinobacteria, and Firmicutes were also identified as an independent negative predictor by multivariate Cox regression.

### Metabolic Pathways and machine learning approach

In a sub-cohort of 37 patients, metagenome pathways were analyzed from the MetaCyc Metabolic Pathway Database. A total of 73 individual Metacyc Pathways were identified. First, Metacyc pathways were classified into 16 “Metacyc SuperPathways” to capture the essential metabolic routes in the bacterial domain (**SFig 5**). The stacked bar chart (**Fig 7A**) shows the distribution of SuperPathways in every patient, where only a limited fraction (11.23%) of high-quality reads were classified as unmapped or unintegrated.

**Figure 7.**
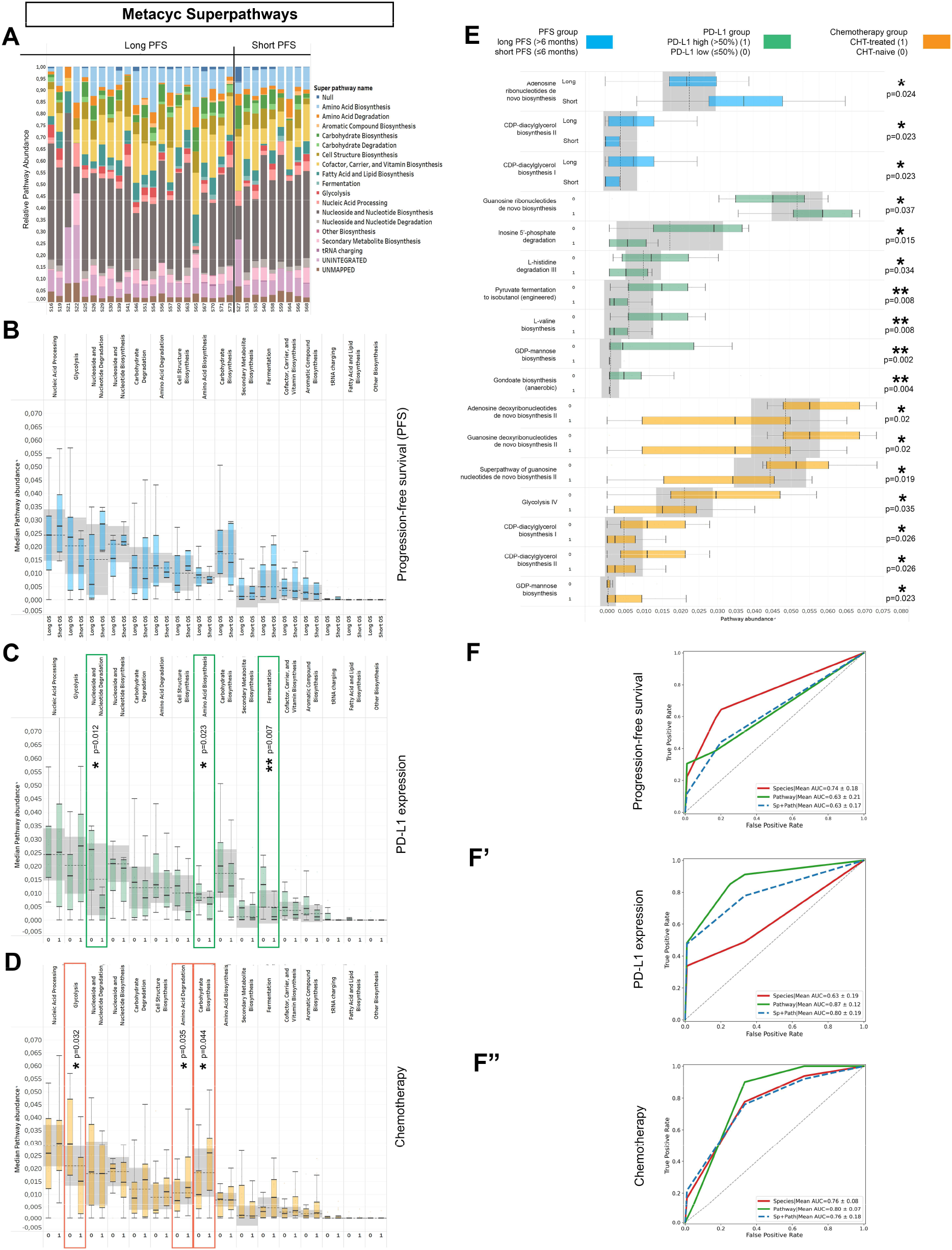
Metagenome Pathways and machine learning approach. Abundance-distribution of Metacyc Superpathways per patient is shown in a stacked bar chart, where X axis shows patient IDs grouped according to PFS and Y axis represents the relative abundance of the correspondent Superpathway, color coded (**A**). Patients were grouped as long vs short PFS, PD-L1-high vs low and CHT-treated vs CHT-naive, as previously described. None of the superpathways showed significant difference according to PFS (**B**), whereas Nucleoside and Nucleotide degradation, Amino acid biosynthesis and Fermentation superpathways were significantly more abundant in PD-L1 low patients (compared to PD-L1 high patients, **C**). Regarding chemotherapy, chemo-naive (0) patients showed significantly increased abundance of the Glycolysis superpathway and significantly decreased abundance of the Amino acid degradation and Carbohydrate biosynthesis superpathways (compared to CHT-treated patients, **D**). Multiple individual Metacyc pathway are differentially abundant between different patient groups, including Adenosine Ribonucleotide de novo Biosynthesis (p=0.024), CDP-diacylglycerol Biosynthesis I-II (p=0.023) according to PFS (**E**); Guanosine Ribonucleotide de novo Biosynthesis (p=0.037), Inosine 5’ Phosphate Degradation (p=0.015) L-histidine Degradation III (p=0.034), Piruvate Fermentation to Isobutanol (p=0.008), L-valine Biosynthesis (p=0.008), GDP-mannose Biosynthesis (p=0.002) and Anaerobic Gondoate Biosynthesis (p=0.004) according to PD-L1 expression (**E**) and GDP-mannose Biosynthesis (p=0.023), Adenosine- and Gianosine Deoxyribonucleotides de novo Biosynthesis II (p=0.02, respectively), Superpathway of Guanosine Nucleotides de novo Biosynthesis II (p=0.019), Glycolisis IV (p=0.035) and CDP-diacylglycerol Biosynthesis I-II (p=0.026, respectively) according to Chemotherapy-regime (**E**). 5-fold cross validation performed on Random Forest (RF) machine learning models show that PFS is best predicted with key species (AUC: 0.74; Recall: 0.64; F1: 0.56, **F**) and PD-L1 expression is best predicted with pathways (AUC: 0.88; Recall: 0.87; F1: 0.82, **F’**). The model fitted fairly to the prediction of first-line (CHT-treated) or subsequent-line (CHT-treated) IT for both key species (AUC: 0.76; Recall: 0.86; F1: 0.82, **F”**) and pathways (AUC: 0.8; Recall: 1; F1: 0.88, **F”**). *Metric data are shown as mean and corresponding standard deviation (SD). Statistical significance *P < 0*.*05; **P < 0*.*01, ***P<0*.*001*.

According to PFS, none of the SuperPathways showed a significant difference in long- vs. short-term survival (**Fig 7B**). In contrast, SuperPathways Nucleoside and Nucleotide degradation (p=0.012), Amino Acid Biosynthesis (p=0.023), and Fermentation (p=0.007) were significantly more abundant in PD-L1-low expressor patients (compared to PD-L1-high expressors, **Fig 7C**). Patients were also assessed according to CHT-treatment: there was an increased abundance of Glycolysis (p=0.032) in CHT-naive patients, whereas there were increased abundances of Amino Acid Degradation (p=0.035) and Carbohydrate Biosynthesis (p=0.044) in CHT-treated patients (**Fig 7D**). Among individual MetaCyc Pathways, patients with short PFS exhibited a significantly increased abundance of Adenosine Ribonucleotide de novo Biosynthesis (p=0.024), whereas CDP-diacylglycerol Biosynthesis I-II are significantly more abundant in patients with long PFS (p=0.023) (**Fig 7E**). There was a more dominant shift in pathway representation according to PD-L1 expression and CHT-regime, where 7-7 pathways showed a significant difference in distinct patient groups, respectively (**Fig 7E**).

Next, a Random Forest (RF) model was built, where 5K-fold cross-validation was used to evaluate the performance of key species and Metacyc Pathway abundances in predicting short vs. long PFS, high vs. low PD-L1 expression, and CHT-treatment or naivety. PFS was best predicted with key species (AUC=0.74), whereas pathways alone (AUC=0.63) or combined with taxonomy (AUC=0.63) gave a poor performance in predicting PFS (**Fig 7F**). In contrast, PD-L1 expression was best predicted with pathways (AUC=0.87) and gave a lower performance for species (AUC=0.63) (**Fig 7F’**). The machine learning algorithm predicted the CHT-regime with a good performance using pathways (AUC=0.8) and with a fair performance using species alone or with species combined with pathways (AUC=0.76, respectively) (**Fig 7F”**). These results suggest that the taxonomic profile is best suited to predict long- or short PFS, while the pathway profile predicts better the PD-L1 phenotype of patients.

## DISCUSSION

The reasons behind the divergent response rates to cancer IT are still poorly understood, and even today, PD-L1 represents the most widely used predictive biomarker in clinics. Therefore, in our study, we analyzed a large-scale, real-life cross-sectional cohort to uncover the theranostic role of the gut microbiome in lung cancer patients. To the best of our knowledge, our study is among the first to provide insights into the specific interactions of the gut microbiome with CHT and IT by analyzing CHT-naïve and CHT-pretreated patients.

Others showed that a higher gut microbiota alpha diversity was associated with increased response rates to ICI therapy and improved PFS [16,17]. In contrast, we found no significant difference in alpha-diversity in any comparison, including PFS, CHT-treatment, and PD-L1 phenotype. However, beta-diversity was significantly different between patients with short and long PFS, as in the case of other NSCLC [16,17] and melanoma studies [9,10,11]. Moreover, we detected a significant gut microbial compositional difference between CHT-treated and CHT-naive patients that underlines the effect of CHT on the gut microbiome. It is noteworthy that bacterial richness and alpha-diversity are parameters that are difficult to assess in the routine clinical practice, whereas detecting or quantifying individual species is cost-effective. Thus, we identified specific indicator species that could predict outcomes with high reliability and moreover, we established a hierarchical model stratifying taxa based on the extent of their predictive role regarding long- or short-term PFS. Importantly, apart from univariate statistical tests, we have also implemented various multivariate statistical models such as Lasso- and Cox proportional hazard regression, to define the predictive relevance of a certain taxon based on strong evidence.

Accordingly, we analyzed the differences regarding major bacterial phyla and found that Firmicutes and Actinobacteria were associated with short PFS. The Firmicutes/Bacteroidetes (F/B) ratio is widely accepted to play an important role in maintaining normal intestinal homeostasis and is impaired in dysbiosis [31,32]. To our knowledge, this is the first NSCLC study to confirm that F/B ratio is significantly increased in patients with short PFS. At the species level, multivariate analysis and validation on an additional cohort unequivocally confirmed the positive predictive role of Alistipes shahii and Alistipes finegoldii and the negative predictive role of Bifidobacterium breve, Streptococcus Salivarius, and Streptococcus vestibularis. A highlighted finding of our analysis is the consistent association of Streptococci with short PFS, where both the genus and multiple species show a stringent association with worse outcomes. In a mixed cohort of epithelial tumors including NSCLC and renal cell carcinoma (RCC) specimens, Routy and colleagues [20] associated taxa Akkermansia muciniphila and Alistipes with better- and Bifidobacteria with worse clinical outcomes, using shotgun metagenomics, similar to the current study. We used the latest version of the KRAKEN reference database that enabled us to discover more extensively the potential predictive role of rare- and low abundance taxa, including an array of Streptococcus species (S. gordonii, S. equinus, and S. anginosus), whose association with negative outcomes was also confirmed by multivariate Cox regression.

Two other NSCLC studies have interpreted the gut microbiome in the context of ICI-response so far. According to Jin et al., Alistipes putredinis, Bifidobacterium longum, and Prevotella copri were associated with better outcomes, while unclassified Ruminococcus was linked with impaired ICI-response [16]. In contrast, Zhang et al. identified Desulfovibrio, Actinomycetales, Bifidobacterium, Anaerostipes, Faecalibacterium, and Alistipes as overrepresented in responder patients and different Fusobacteria in non-responders [17]. However, none of these studies interpreted the taxonomic profiles at the species level. Both research groups assessed the gut microbiome in an East-Asian patient cohort, whose baseline gut commensal flora significantly differed from the Caucasian [33]. Other possible explanations for the discrepancies include the different sample collection frame times, reference databases (Metaphlan vs. KRAKEN), and study endpoints. It is important to highlight that previous NSCLC studies interpreting alpha-diversity and taxonomic profiles according to ICI-response [16,17] used 16S rRNA sequencing works with distinct reference datasets and limited taxonomical scope compared to shotgun metagenomics.

Next, we showed that the network of bacterial communities is more interconnected in patients with long PFS, with stronger correlations between taxa. S. vestibularis, B. uniformis, B. faecalis, A. shahii, and B. visceriola were the central hubs in the community network. Our study is unique in terms of first interpreting taxonomic differences according to CHT treatment or naivety in the context of ICI efficacy. While multiple taxa show differential abundance between the CHT-naive and CHT-treated population, CHT-treatment itself does not shift the microbiome into a more short- or long PFS-like profile. The fact that the profile of differentially abundant taxa (in the context of PFS on ICI) in CHT-treated patients and the whole cohort significantly overlap underpins the principle that CHT pretreatment is not detrimental to the microbiome concerning ICI efficacy. CHT-pretreated patients exhibit slightly worse outcomes (vs. CHT-naive patients), possibly due to their tumor’s lower PD-L1 expression. Further studies are needed to confirm these findings.

We revealed that treatment toxicity is not associated with an extensive shift in the microbiome, Prevotella dentalis being the only species with a marked increase in patients with ICI-related trAEs. There is still a contradiction about whether AB treatment significantly affects ICI efficacy. While Pinato and colleagues claim that AB therapy reduced response rates prior, but not during ICI-treatment [26], others reported the opposite or the lack of significance in multivariate analysis [34,35]. Our findings indicate a noticeable change in microbial signature, with the strongest alteration experienced in Eggerthellas; specifically, E. lentha, and no taxa associated with short PFS show significantly increased abundance in the AB-treated group. So far, only preclinical studies have reported the shift in the intestinal microbiome due to systemic corticosteroid treatment [27,36]. We found only a minor bacterial conversion in steroid-treated patients with an increased abundance of Proteobacteria, including Escherichia and E.coli. In contrast, we observed a prominent increase of short-PFS-associated Streptococcus species in patients treated with antacids. This is supported by findings of a recent meta-analysis claiming that PPI medication disrupts the gut microbiome and worsens outcomes of ICI therapy [37].

Analyzing functional metabolic profiles of the gut microbiome is an outstanding advantage of shotgun metagenomics compared to 16S RNA sequencing. SuperPathways Nucleoside and Nucleotide degradation, Amino Acid Biosynthesis, and Fermentation were significantly increased in PD-L1 low patients, whereas Amino Acid Degradation and Carbohydrate Biosynthesis were significantly increased in CHT-treated patients, and Glycolysis was significantly increased in CHT-naive patients. When analyzing taxonomic profiles, we revealed that Euryarchaeota phylum [38] and its prominent species Methanobrevibacter smithii showed a strong association with PD-L1 expression being overrepresented in low expressors, which might be connected to the fact that there was an increased rate of fermentation [38] in PD-L1-low patients. Moreover, the machine learning approach with the Random Forest model confirmed that predicting PD-L1 phenotype has a higher accuracy using pathways, whereas taxonomic profiles performed better when predicting short vs. long PFS.

The strengths of our study include the short turnaround time of sample collection, the careful attention to related methods using the latest and most comprehensive databases, and the measurement of multiple confounding factors associated with host genetics and exposures. PFS-associated taxa are presented in a hierarchical manner supported by multivariate testing, a novel approach in our research. Limitations of our study include that we cannot assess whether altered microbiota contribute to or exist as a consequence of disease. However, this is a widespread issue in the field currently. Therefore, we are cautious with interpreting our results and encourage further studies with a greater sample size. Additionally, we did not have next-generation sequencing performed on patients’ tumor tissue samples to be able to discern the potential impact of tumor mutation burden (TMB). However, unlike PD-L1, TMB has not been confirmed to be predictive for OS in NSCLC [39].

## CONCLUSION

Our study shows the complexity of the gut microbiome, a highly diverse system, in NSCLC. We were able to define an outcome-related common gut microbial signature with internal cross-validation and using an additional cohort. Multiple Streptococcus and Bifidobacteria species showed a stringent association with impaired ICI-efficacy, whereas the presence of Alistipes and Barnesiella was linked to better outcomes. The machine learning approach revealed that the PD-L1 phenotype is best predicted from metabolic pathways, while the taxonomic profile is more suited to predict outcomes. To our knowledge, this is the first comprehensive data in NSCLC using metagenomic visualization tools to extract knowledge efficiently to support further studies in the field.

## Data Availability

Data are available on reasonable request. Data relevant to the study are included in Supplementary material or available on reasonable request to the correspondence author.

## CRediT Authorship Contribution Statement

**David Dora:** Conceptualization, formal analysis, investigation, data curation, visualization, supervision, funding acquisition, writing-original draft, writing—review and editing

**Balazs Ligeti:** data curation, formal analysis, investigation, visualization, funding acquisition

**Tamas Kovacs:** formal analysis, investigation, visualization

**Peter Revisnyei:** data curation, formal analysis, visualization

**Gabrielly Galffy:** data curation, resources

**Edit Dulka**: data curation, resources

**Daniel Krizsán:** data curation, formal analysis

**Regina Kalcsevszki:** data curation, formal analysis

**Zsolt Megyesfalvi:** formal analysis, validation, writing—review and editing

**Balazs Dome:** investigation, validation, supervision, resources, funding acquisition, writing— review and editing

**Glenn J. Weiss:** validation, supervision, resources, writing—review and editing

**Zoltan Lohinai:** Conceptualization, investigation, supervision, resources, funding acquisition, writing-original draft, writing—review and editing

## ACKNOWLEDGEMENTS

BD and ZM acknowledge funding from the Hungarian National Research, Development and Innovation Office (KH130356 and KKP126790 to BD; 2020-1.1.6-JÖVŐ and TKP2021-EGA-33 to BD and ZM). BD was also supported by the Austrian Science Fund (FWF I3522, FWF I3977 and I4677). ZM was supported by the UNKP–20–3 and UNKP–21–3 New National Excellence Program of the Ministry for Innovation and Technology of Hungary, and by the Hungarian Respiratory Society (MPA #2020). DD and BL acknowledge funding from the Hungarian National Research, Development and Innovation Office (142287 to DD; 138055 to BL). BL was also supported by the Thematic Excellence Program (TKP2020-NKA-11).

