## Supplementary material for "Non-Small Cell Lung Cancer Patients Treated with Anti-PD1 Immunotherapy Show Distinct Microbial Signatures and Metabolic Pathways According to Clinical Outcomes"

### SUPPLEMENTAL METHODS

#### Shotgun metagenome pipeline

After signing informed consent, patients were included with baseline stool samples defined as collected within seven days, either before or after the first IV dose of ICI administration. On the day of collection, all samples were placed in the -80°C freezer until they were isolated and sequenced. For whole DNA extraction, we used 100mg stool sample in ZR Bashing Bead Lysis Tubes with ZymoBIOMICS 96 MagBead DNA kit, by continuous bead beating for 40 minutes, and centrifuging the lysate for 1 min. at 10,000 x g. 200 µl supernatant was mixed with 25 µl ZymoBIOMICS™ MagBinding Beads, then shaked for 10 minutes. After placing the tubes on magnetic rack and removing the supernatant, 500 µl ZymoBIOMICS™ MagBinding Buffer was added each sample and mixed for 1 minute. The beads were pelleted, and washed 2 times with 500 µl of ZymoBIOMICS™ MagWash 1 and 900 µl ZymoBIOMICS™ MagWash 2 respectively for 1 min. The beads were dried at 55°C for 10 min. then eluted in 50 µl RNAse/DNAse free water. The DNA concentration was measured with Qubit fluorimeter.

From each sample 65 ng was used as input for library preparation by KAPA HyperPlus kit as per the manufacturer instructions, with size selection for ~200bp peak fragment size (TapeStation 2200, High Sensitivity D1000 ScreenTape®). The samples were sequenced on NextSeq500 platform, 2x150bp with ~10M read pairs.

#### Taxonomic assignment

The reads were adaptor-trimmed and quality-filtered for a minimum mean Q-score of 30. Quality check was performed using fastQC, to remove the adapter regions, low-quality reads, and human DNA contaminations (bwa (version 0.7.4-r385) passing per sequence quality score, per base N content and adapter content [40].

For taxonomic assignment Kraken2 was used (version 2.0.8) [41] with the MiniKraken2 database. The result files were merged into a data matrix with KrakenTools (v1.2) combine_kreports.py script. The read counts were normalised by rarefaction using the smallest sample as minimum depth with inclusion criteria of min. 1 read in min. 1 sample per taxa. A significant proportion of the reads had fallen in the unclassified bin (mean=0.58, SD=0.086). The results were stratified by taxa for statistical analysis. In further analyses, only taxa within domains Bacteria and Archea were included, we excluded all viral and eukaryotic taxonomic units. Rare (present maximum 10% in all samples) and low abundance (support of less than 0.01% abundance) taxa were discarded from the subsequent analysis. After the filtering process, a Bayesian-Multiplicative replacement of zeros was carried out using the z Composition R package, which was followed by central log-ratio (CLR) transformation of count and ratio values as implemented in scikit-bio. In CLR-transformation, sample vectors undergo a transformation based on the logarithm of the ratio between the individual elements and the geometric mean of the vector. The Shannon index was used for measure of alpha diversities, which quantifies the entropy of the distributions of taxa and functional proportions.

#### Microbial networks

Interaction networks from bacterial taxa (phylum, genus, species) were built in three different groups (all patients, patients with short PFS- and long PFS) with Cytoscape [42]. We filtered the edges to keep only **significant (p<0.01) and at least moderately correlated interactions r≥|0.4|. We removed all self-loops and duplicated edges. Consensus (intersections) and differential networks (subtractions) were made by Cytoscape's built-in merge networks function. The differential networks between long PFS- and short PFS groups in each taxa were made by the Diffany plugin [43] of the same software package. Long PFS patients were defined as reference group for all comparison networks.

**Pathway analyses**

Metagenomic functional profiles using MetaCyc Pathways and Superpathways were available in a subset of Discovery cohort for n=37 patients. The quality of raw and trimmed reads was assessed with FastQC and MultiQC. The low-quality sequences were filtered and trimmed by Trimmomatic and only sequences with minimal length of 36 and low quality base calls were discarded (phred score <30). The host contamination (reads aligning to human reference genome (bowtie2 v2.4.2 [44], GRCh38)) were discarded. Pathway abundance and other molecular functional profiles (such as SuperPathways) were estimated with MetaPhlan3 and HUMAnN3 pipeline [45].

**Shotgun Metagenome pipeline for validation cohort**

For the Validation cohort (n=60), high-quality reads were taxonomically annotated using MetaPhlAn2 (version 2.7.7) with the default settings. The differentially abundant taxa were found on the unrarefied relative abundance data using the Wald test implemented in the R package DESeq2 v1.22.2 [46], and the statistical significance was filtered with p<0.05 unless otherwise stated, as previously described [47,48].

**Random forest model and machine learning approach**

Multiple random forest model with stratified five-fold-cross-validation were created for binary classification, using the scikit-learn (1.1.2) python (3.10.6) package. PFS (long vs short), PD-L1 (high vs low) and Chemotherapy (CHT-naïve vs treated) were used separately as target variables. The individual models were trained on key bacterial species (**Fig 3A**) and pathway datasets (**Fig 7**). Both datasets were utilized separately and combined too. The models' performance was evaluated using the area under the ROC curve (AUC) values.

**PD-L1 Immunohistochemistry (IHC)**

Tumor samples from advanced-stage NSCLC patients were available for PD-L1 IHC for n=50 patients retrieved by lung biopsy. For IHC staining, four-µm-sections were cut from each formalin-fixed-paraffin-embedded (FFPE) blocks. Staining was carried out on a Leica Bond RX autostainer using rabbit monoclonal antibody for PD-L1 diluted 1:300 (CST, cat: 13684S), and rabbit monoclonal antibody for PD-1 diluted 1:400 (CST, cat: 86163). Slides were stained with the Bond Polymer Refine Detection kit (#DS9800) and Leica IHC Protocol F, and epitope retrieval was carried out for twenty minutes at low pH. Slides were cleared and dehydrated on a Tissue-Tek Prisma platform before being coverslipped using a Tissue-Tek Film coverslipper. PD-L1 expression was evaluated by an expert histopathologist. Patients were stratified according to high (≥50$^{th}$ percentile) or low (<50$^{th}$ percentile) expression.

**Statistical analyses**

The visualization of the microbiome's taxonomic and pathway composition (beta-diversity) was carried out with Uniform Manifold Approximation and Projection (UMAP) using the CLR values as input matrix (scikit-learn v0.24) [49]. The compositional similarities between different groups were investigated with permutational analysis of variance (PERMANOVA) and the differential abundance testing was done using the Wilcoxon rank-sum test. The associations between CLR-normalized abundances of taxa were investigated with Spearman's rank correlation for network analyses. Survival analysis was carried out using Kaplan-Meier (KM) curves, and survival curves were compared using the log-rank test. Cut-offs for KM curves and specificity/sensitivity values for taxa were defined by Receiver Operating Characteristic (ROC) curve analysis with using the binary outcomes of short vs long PFS.

Least absolute shrinkage and selection operator (Lasso) regression was used to select the most predictive markers from our high-dimensional data and reduce the interaction between markers to avoid overfitting. Lasso regression is distinguished by variable selection and complexity regularization when fitting the generalized linear model. The optimal hyperparameters were selected via 5-fold cross-validation procedure using ROC-AUC as target metric. To identify relevant predictor factors, Cox-proportional hazard regression was performed. The analysis was two-sided, with a significance threshold of=0.05. In multivariate Cox regression, the backwards elimination method was used, where parameters ($p<0.1$) were included. Harrel's C-index was calculated to assess the quality of fit of our multivariate model that performed above 0.7 (fair) in all analyses.

### SUPPLEMENTAL FIGURES

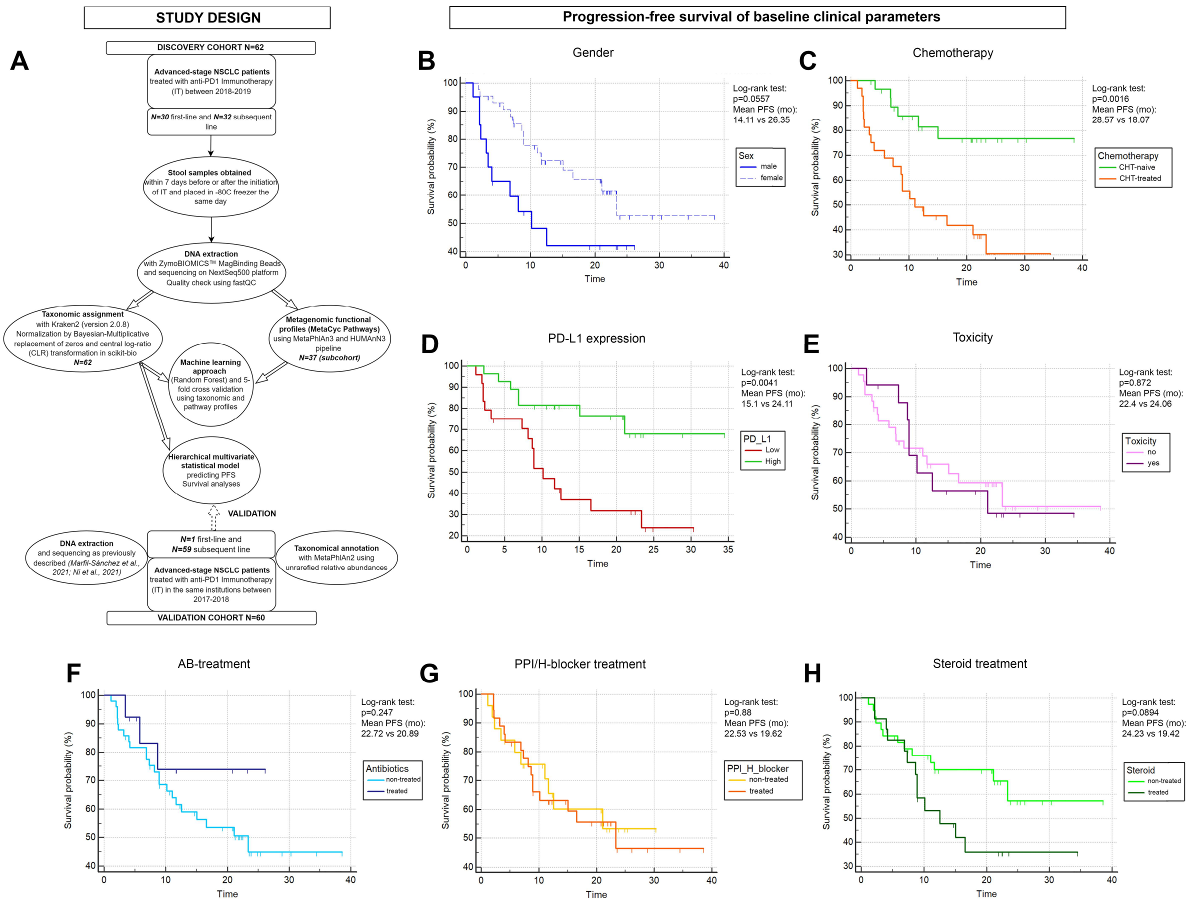

**SFig 1. Flowchart of study design and KM analysis of baseline clinical parameters.** Flowchart demonstrates study design, cohorts and experimental procedures (**A**). PFS was assessed for baseline clinical parameters in KM curves for gender (**B**), Chemotherapy-regime (CHT-naïve vs CHT-treated, **C**), PD-L1 expression low (<50%) vs high (≥50%), **D**) and Treatment-toxicity (**E**). Medications prior IT were also assessed, including AB-treatment (**F**), Antacid medication (**G**) and Steroid treatment (**H**). Female patients exhibited a trend towards increased progression-free survival (PFS) compared to males, p=0.0557, **B**). CHT-naïve- and PD-L1-high patients showed significantly increased PFS (compared CHT-treated (p=0.0016, **C**) and PD-L1 low patients (p=0.0041, **D**), respectively. There was no significant difference in PFS according to Treatment-toxicity (p=0.872, **E**), AB-treatment (0.247, **F**), PPI/H-blocker medication (p=0.88, **G**) and steroid treatment (p=0.0894, **H**).

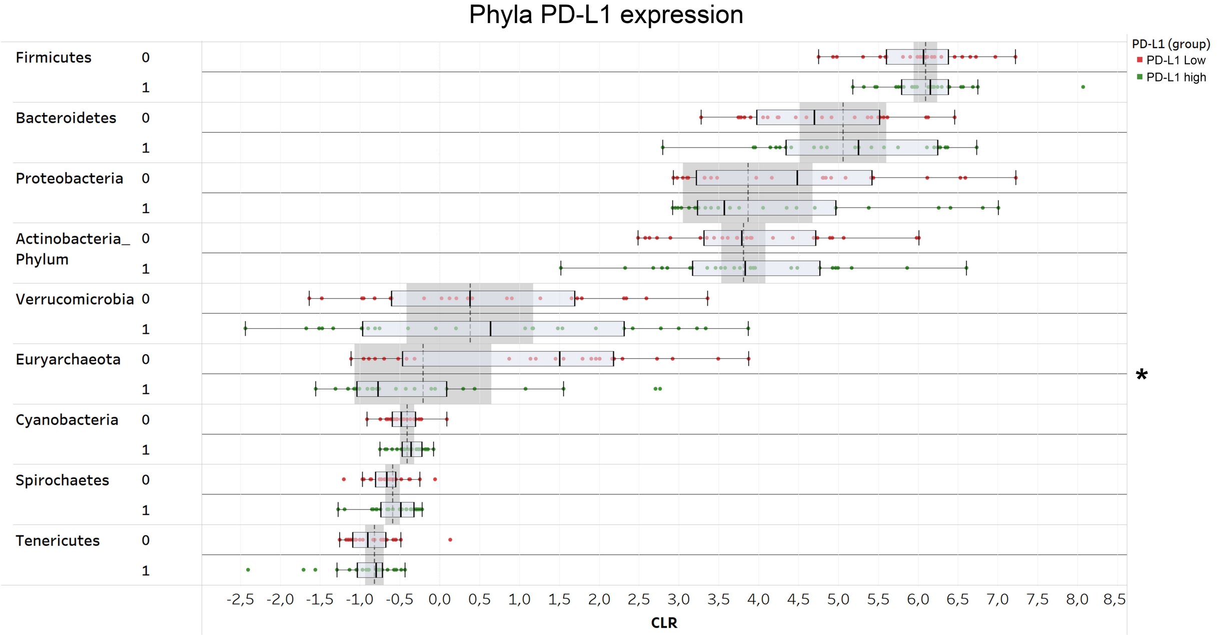


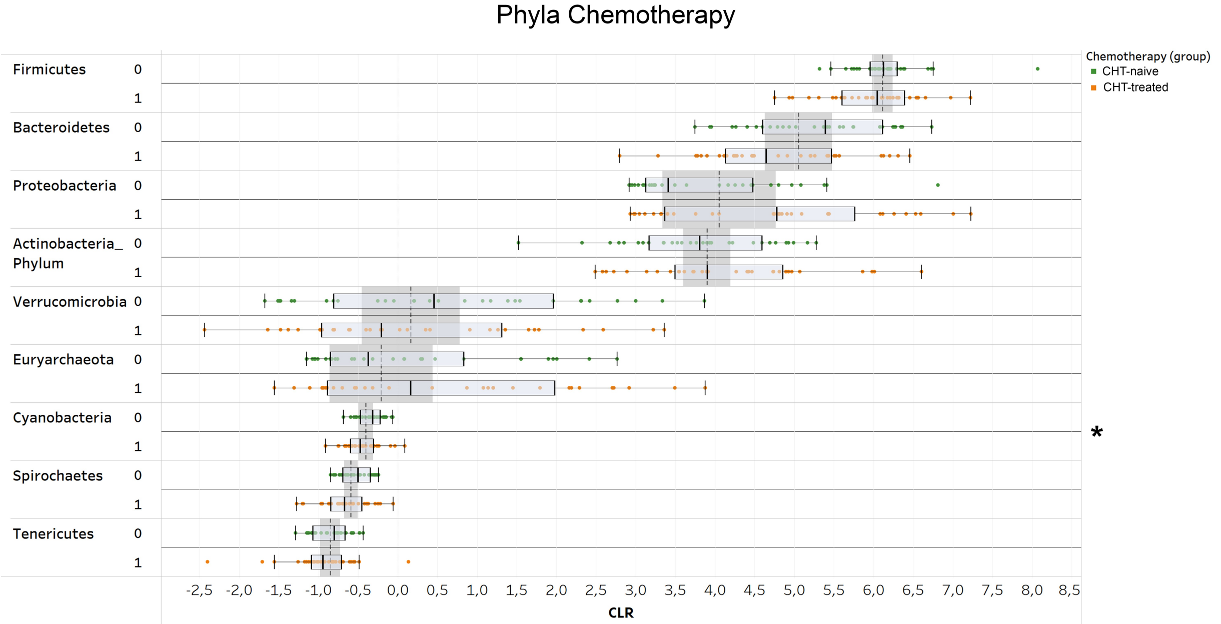


**SFig 2. Abundance of major phyla according to PD-L1 expression, and CHT.** Euryarcheota are significantly more abundant in PD-L1 low patients (compared to PD-L1 high). Cyanobacteria are significantly more abundant in CHT-naive patients. There were no significant differences according to

sex. *Metric data are shown as mean and corresponding standard deviation (SD). Statistical significance *P < 0.05; **P < 0.01, ***P<0.001.*

**A**

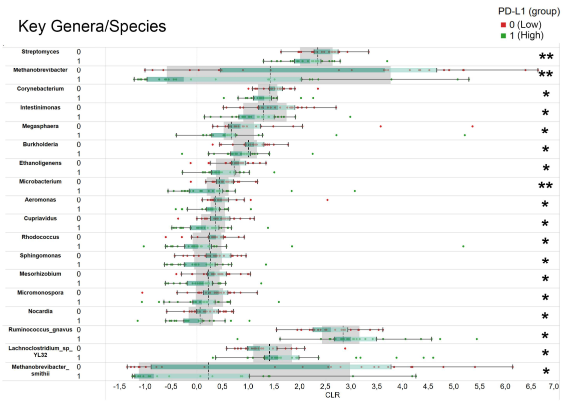


**B**

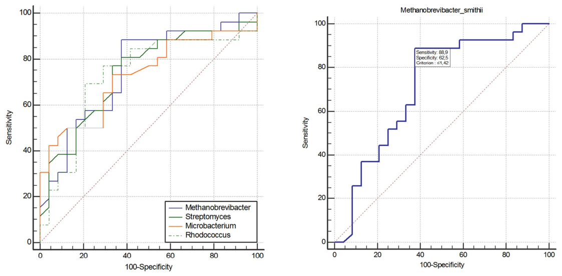


| GENUS | ROC AUC | 95% CI | p-value | Sensitivity | Specificity |
|---|---|---|---|---|---|
| Methanobrevibacter | 0,752 | 0,615 to 0,890 | 0,0003 | 88,46 | 62,5 |
| Streptomyces | 0,74 | 0,599 to 0,881 | 0,0009 | 76,92 | 66,67 |
| Microbacterium | 0,727 | 0,583 to 0,870 | 0,0019 | 73,08 | 66,67 |
| Rhodococcus | 0,728 | 0,579 to 0,878 | 0,0027 | 69,23 | 79,17 |
| **SPECIES** | | | | | |
| Methanobrevibacter smithii | 0,714 | 0,564 to 0,864 | 0,0053 | 88,89 | 62,5 |

**C**

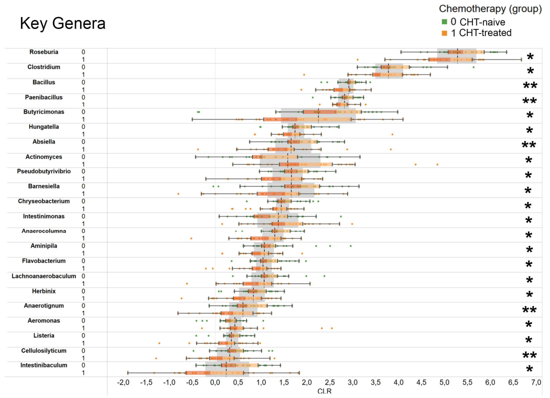


**D**

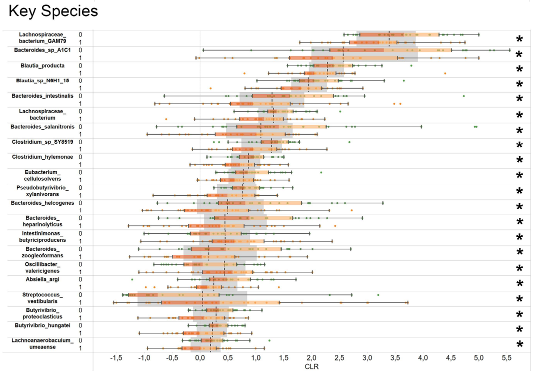


**SFig 3. Taxonomy of genera and species according to PD-L1 expression and Line of IT.** Abundances of taxa in PD-L1 high expressors (≥50% (1)) vs PD-L1 low expressors (<50% (0)) are shown in panel **A.** Only genera (CLR>0) and species (CLR>-1), where there was a significant difference in abundance between PD-L1-high and low patients according to Wilcoxon ranksum test (p<0.05) are listed. ROC curves of genera (with at least 0.7 AUC of predicting PD-L1 phenotype) and species M. smithii with are shown in panel **B.** Sub-table in panel **B** shows ROC AUC, 95% CI, p-value of ROC analysis, sensitivity and specificity to predict PD-L1 phenotype in key genera and species (AUC>0.7). Abundances of genera and species in CHT-naive (0) vs CHT-treated (1) patients are shown in panel **C** and **D**, respectively. Only genera (CLR>0) and species (CLR>-1), where there was a significant difference in abundance between PD-L1-high and low patients according to Wilcoxon ranksum test (p<0.05) are listed. *Metric data are shown as mean and corresponding standard deviation (SD). Statistical significance *P < 0.05; **P < 0.01, ***P<0.001.*

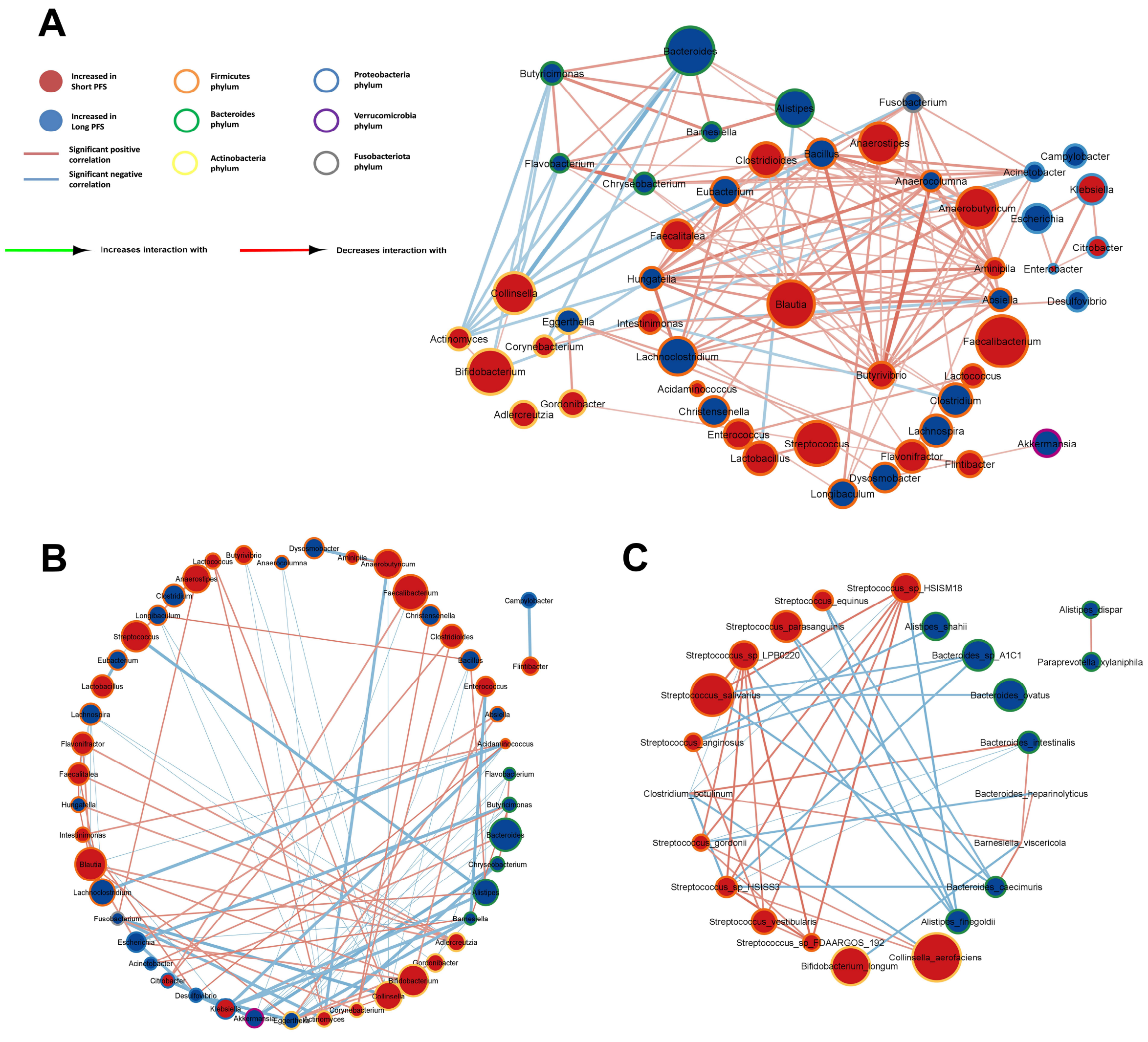


**SFig 4. Whole network for all genera and differential networks for all genera and key species.** Network generated from all major genera (CLR>0) in the whole cohort with legends are shown in panel **A**. Differential network in the same setup shows connections occurring only among patients with short PFS, but not among patients with long PFS (**B**). The same is shown in panel **C** for key species.

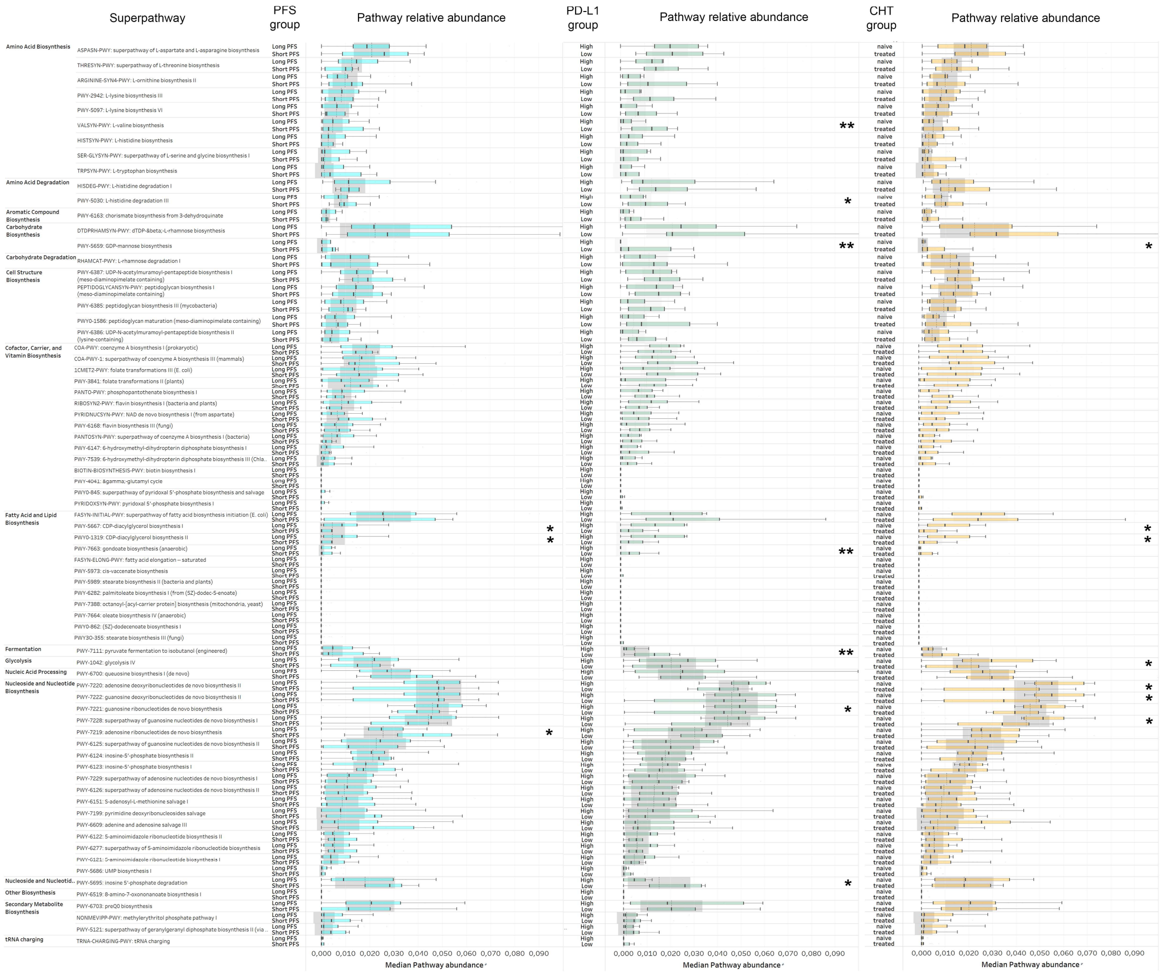

**SFig 5. All pathways arranged according to their Superpathway class in long- vs short PFS, PD-L1 low- vs high and in CHT-naive- vs treated patients.**

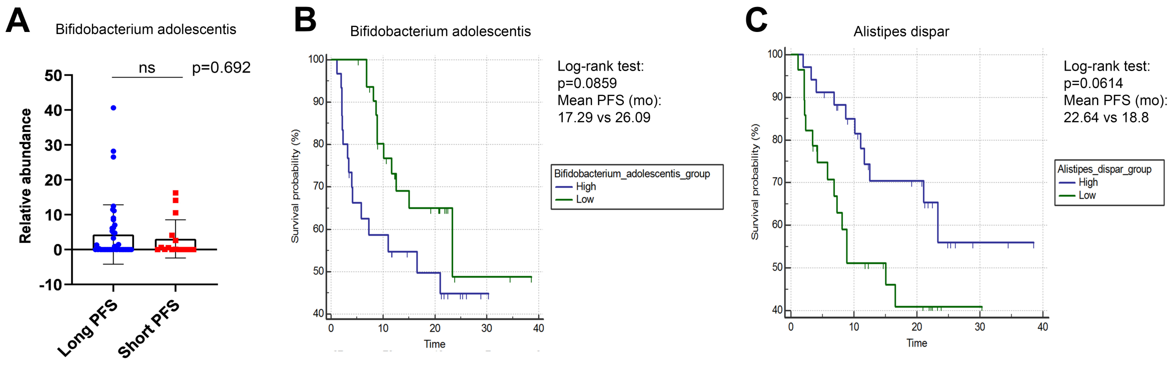


**SFig 6. Non-significant KMs and validation.** In the validation cohort, abundance of B. adolescentis show no significant difference in long- vs short PFS patients (p=0.692, **A**). B. adolescentis (p=0.0859, **B**) and A. dispar (p=0.0614, **C**) show no significant difference according to KM analysis.

### SUPPLEMENTAL TABLES

**STable 1.** Immunotherapy drugs administered to patients

| | Pembrolizumab N=29 (47%) | Nivolumab N=22 (35%) | Durvalumab N=7 (12%) | Atezolizumab N=4 (6%) | p-value |
|---|---|---|---|---|---|
| Gender (male) | 20/29 | 9/22 | 5/7 | 0/4 | 0.021* |
| Response at 3 months (R) | 29/29 | 18/22 | 7/7 | 2/2 | 0.051 |
| PFS group (long) | 24/29 | 15/22 | 6/7 | 2/4 | 0.573 |
| Chemotherapy (treated) | 6/29 | 22/22 | 0/7 | 4/4 | <0.001*** |
| PD-L1 IHC (>50%) | 26/29 | 0/22 | 0/7 | 1/4 | <0.001*** |
| Smoking (pack-year) | 40.88 | 36.89 | 43.57 | 47.83 | 0.948 |
| ICI Toxicity (present) | 6/29 | 7/22 | 1/7 | 1/4 | 0.709 |
| Antibiotics (pre-ICI) | 7/29 | 3/22 | 2/7 | 0/4 | 0.581 |
| Steroids (pre-ICI) | 10/29 | 9/22 | 1/7 | 2/4 | 0.564 |
| Antacids (pre-ICI) | 17/29 | 12/22 | 4/7 | 2/4 | 0.984 |

**STable 2.** Comparison of first-line (CHT-naive) vs subsequent-line IT patients

| | CHT-naive patients N=30 (48%) | CHT-treated patients N=32 (52%) | p-value |
|---|---|---|---|
| Age (mean) | 58.72 | 60.68 | 0.134 |
| Gender (male) | 9 (30%) | 11 (34%) | 0.789 |
| Initial Response (R) at month 3 | 30 (100%) | 24 (75%) | 0.004** |
| PFS group (long) | 25 (83%) | 22 (69%) | 0.111 |
| PD-L1 IHC (>50%) | 20 (66%); n/a = 6 (20%) | 6 (19%); n/a = 6 (19%) | <0.001*** |
| Smoking (pack-year) | 37.27 | 42.76 | 0.442 |
| ICI Toxicity (present) | 5 (17%) | 12 (37%) | 0.089 |
| Antibiotics (pre-ICI) | 9 (30%) | 4 (12%) | 0.122 |
| Steroids (pre-ICI) | 7 (23%) | 16 (50%) | 0.063 |
| Antacids (pre-ICI) | 17 (56%) | 19 (59%) | >0.99 |

**STable 3.** Clinicopathological characteristics of Validation cohort. *P-values are indicated for Fischer's exact test, Mann-Whitney test or Welch's test in case of normality.*

| | Long PFS N=44 (73%) | Short PFS N=16 (27%) | p-value |
|---|---|---|---|
| Age (mean) | 60.12 | 62.41 | 0.121 |
| Gender (male) | 38.63% (17) | 37.5% (6) | >0.99 |
| Response at 3 months (R) | 100% (44) | 31.25% (5) | <0.001 *** |
| PFS (months, median) | 12.64 | 3.67 | <0.001 *** |
| Chemotherapy (treated) | 97.73% (43) | 100% (16) | >0.99 |
| PD-L1 IHC (>50%) | 20.45% (9) | 12.5% (2) | 0.714 |

**Stable 4.** Comparison of clinicopathological characteristics between Discovery and Validation cohorts. *P-values are indicated for Fischer's exact test, Mann-Whitney test or Welch's test in case of normality.*

| | Discovery cohort | Validation cohort | p-value |
|---|---|---|---|
| Age (mean) | 60.05 | 61.23 | 0.104 |
| Gender (male) | 32.25% (20) | 38.33% (23) | 0.57 |
| Response (R) | 82.25% (51) | 81.66% (49) | 0.823 |
| PFS (months, median) | 14.67 | 12.53 | 0.25 |
| Chemotherapy (treated) | 51.61% (32) | 98.33% (59) | <0.001 *** |
| PD-L1 IHC (>50%) | 51.61% (32) | 18.33% (11) | 0.001 ** |

**STable 5a.** Univariate Cox regression, baseline clinicopathological parameters

| **Covariate** | **b** | **SE** | **Wald** | **P** | **Exp(b)** | **95% CI of Exp(b)** |
|---|---|---|---|---|---|---|
| Gender=0 | 0.743 | 0.398 | 3.487 | 0.061 | 2.103 | 0.967 to 4.57 |
| **Chemotherapy=0** | -1.364 | 0.465 | 8.571 | **0.003** | 0.255 | 0.103 to 0.634 |
| **PD-L1 IHC=0** | 1.228 | 0.455 | 7.262 | **0.007** | 3.415 | 1.404 to 8.307 |
| Toxicity=1 | 0.068 | 0.429 | 0.025 | 0.872 | 1.071 | 0.463 to 2.473 |
| Antibiotics=1 | -0.695 | 0.614 | 1.282 | 0.257 | 0.498 | 0.15 to 1.652 |
| Antacids=0 | -0.06 | 0.403 | 0.0225 | 0.88 | 0.941 | 0.428 to 2.066 |
| Steroids=1 | 0.657 | 0.394 | 2.776 | 0.095 | 1.93 | 0.89 to 4.169 |

**STable 5b.** Multivariate Cox regression, baseline clinicopathological parameters. Backwards elimination (p>0.1).

| Covariate | b | SE | Wald | P | Exp(b) | 95% CI of Exp(b) |
|---|---|---|---|---|---|---|
| **PD-L1 IHC=0** | 1.351 | 0.481 | 7.887 | **0.0050** | 3.861 | 1.511 to 9.867 |
| Gender=0 | | | | >0.1 | | |
| Chemotherapy=0 | | | | >0.1 | | |
| ICI Toxicity=1 | | | | >0.1 | | |
| Antacids=0 | | | | >0.1 | | |
| Steroids=1 | | | | >0.1 | | |

**STable 6. Top PFS-associated taxa in CHT-treated patients**

| tax_level | taxa_id | W p_value | W pos_median | W neg_median |
|---|---|---|---|---|
| s | Alistipes_shahii | 0.0155 | 2.5943 | 1.2107 |
| s | Streptococcus_salivarius | 0.0175 | 1.7693 | 3.3997 |
| s | Escherichia_coli | 0.0278 | 4.406 | 1.3479 |
| s | Streptococcus_vestibularis | 0.0347 | 0.0799 | 1.6127 |
| s | Ruminococcus_bicirculans | 0.0347 | 1.9031 | 3.5197 |
| s | Bifidobacterium_adolescenti | 0.0386 | 1.2687 | 3.2653 |

**STable 7. Characteristics of bacterial clusters**

| | **Bacterial cluster** | **α** | **β** | **γ** | **δ** |
|---|---|---|---|---|---|
| | **Cluster Description** | Beneficial Bacteroides | Detrimental Streptococci | Other Beneficial Bacteroidetes | Other Detrimental Bacteria |
| **Abundance in patient cluster** | Cluster A1a | High | Low | Moderate | Moderate |
| | Cluster A1b | High | Low | High | Low |
| | Cluster A2 | High | Moderate | High | Low to Moderate |
| | Cluster B1 | Low to moderate | High | Low | High |
| | Cluster B2 | Low to moderate | Moderate | Low to Moderate | High |

**STable 8. Wilcoxon test results and all ROC-analysis results**

| Taxon | W p_value | W pos_median | W neg_median | Lasso coeff n | ROC AUC | 95% CI | p-value | Sensitivity | Specificity |
|---|---|---|---|---|---|---|---|---|---|
| **GENUS** | | | | | | | | | |
| Barnesiella | 0.0014 | 1.9514 | 1.3947 | 1.8581 | 0.77 | 0.646 to 0.894 | <0.0001 | 87.5 | 63.83 |
| Butyricimonas | 0.0025 | 2.6414 | 1.5548 | 1.0016 | 0.762 | 0.628 to 0.896 | 0.0001 | 75 | 76.6 |
| Bifidobacterium | 0.0028 | 5.192 | 6.3538 | | 0.749 | 0.613 to 0.886 | 0.0003 | 68.75 | 74.47 |
| Streptococcus | 0.0171 | 4.7325 | 5.7114 | | 0.7 | 0.547 to 0.853 | 0.0104 | 37.5 | 95.74 |
| Alistipes | 0.0186 | 5.6296 | 4.5803 | | 0.694 | | | | |
| Methanosphaera | 0.0202 | -0.9985 | 0.058 | 1.2317 | 0.699 | 0.554 to 0.845 | 0.0071 | 75 | 72.34 |
| Buchnera | 0.0211 | -0.3933 | -0.5809 | | 0.704 | 0.559 to 0.849 | 0.0057 | 56.25 | 80.85 |
| Collinsella | 0.0249 | 4.5016 | 5.227 | | 0.686 | | | | |
| Actinomyces | 0.0317 | 1.4640 | 1.95 | | 0.672 | | | | |
| Paraprevotella | 0.0317 | 2.0788 | 1.2787 | 1.1755 | 0.684 | | | | |
| Helicobacter | 0.0330 | -0.0137 | -0.2544 | | 0.689 | | | | |
| Spiroplasma | 0.0343 | 0.1617 | -0.047 | | 0.684 | | | | |
| Lactobacillus | 0.0432 | 3.312 | 3.8056 | | 0.674 | | | | |

| | | | | | | | | | |
|---|---|---|---|---|---|---|---|---|---|
| Sutterella | 0.2759 | -0.3999 | 0.0213 | 0.9232 | 0.597 | | | | |
| Adlercreutzia | 0.1217 | 1.2177 | 2.5225 | 0.7513 | 0.63 | | | | |
| **PHYLUM** | | | | | | | | | |
| Actinobacteria | 0.0034 | 3.6125 | 4.7046 | 0.5961 | 0.735 | 0.608 to 0.838 | 0.0009 | 68.75 | 74.47 |
| Firmicutes | 0.0220 | 5.9358 | 6.2139 | 0.2233 | 0.711 | 0.583 to 0.818 | 0.0067 | 62.5 | 78.72 |
| Bacteroidetes | 0.1899 | 5.1456 | 4.7788 | 0.1939 | 0.635 | | | | |
| Euryarchaeota | 0.7883 | -0.582 | 0.348 | 0.1463 | 0.513 | | | | |
| Proteobacteria | 0.6470 | 4.1912 | 3.7441 | 0.1398 | 0.543 | | | | |
| Verrucomicrobia | 0.4390 | 0.0486 | -0.4871 | 0.0665 | 0.57 | | | | |
| **SPECIES** | | | | | | | | | |
| Alistipes_shahii | 0.0003 | 2.6453 | 1.267 | 1.4665 | 0.81 | 0.691 to 0.898 | <0.0001 | 87.5 | 74.47 |
| Barnesiella_visc ericola | 0.0007 | 1.0819 | 0.2734 | 0.4796 | 0.787 | 0.666 to 0.880 | <0.0001 | 75 | 74.47 |
| Butyricimonas_fa ecalis | 0.0014 | 1.7255 | 0.5503 | | 0.771 | 0.648 to 0.868 | <0.0001 | 68.75 | 82.98 |
| Streptococcus_s alivarius | 0.0029 | 1.6534 | 3.4884 | 0.5814 | 0.755 | 0.631 to 0.855 | 0.0001 | 87.5 | 53.19 |
| Bacteroides_sp_ A1C1 | 0.0031 | 3.5467 | 2.112 | | 0.735 | 0.609 to 0.839 | 0.0007 | 93.75 | 51.06 |

| | | | | | | | | | |
|---|---|---|---|---|---|---|---|---|---|
| Alistipes_finegoldii | 0.0040 | 2.1055 | 1.2275 | 1.0919 | 0.741 | 0.616 to 0.844 | 0.0002 | 93.75 | 57.45 |
| Streptococcus_vestibularis | 0.0057 | -0.4302 | 0.9065 | | 0.734 | 0.608 to 0.837 | 0.0009 | 81.25 | 59.57 |
| Bifidobacterium_adolescentis | 0.0060 | 0.4987 | 3.2653 | 0.4185 | 0.734 | 0.608 to 0.837 | 0.0004 | 87.5 | 63.83 |
| Bifidobacterium_breve | 0.0079 | -0.1853 | 0.5889 | | 0.727 | 0.600 to 0.831 | 0.0025 | 68.75 | 72.34 |
| Streptococcus_parasanguinis | 0.0100 | 0.7426 | 1.7425 | | 0.717 | 0.590 to 0.824 | 0.0022 | 62.5 | 76.6 |
| Alistipes_dispar | 0.0126 | 1.2804 | 0.3676 | 0.9224 | 0.707 | 0.579 to 0.815 | 0.0028 | 75 | 63.83 |
| Bacteroides_caecimuris | 0.0163 | 1.5349 | 0.5408 | | 0.691 | | | | |
| Bifidobacterium_longum | 0.0171 | 2.2285 | 3.7577 | | 0.699 | | | | |
| Bacteroides_ovatus | 0.0186 | 3.1615 | 2.2042 | 0.2265 | 0.697 | | | | |
| Streptococcus_sp_HSISM1 | 0.0202 | -0.0133 | 1.3179 | | 0.699 | | | | |
| Bacteroides_intestinalis | 0.0220 | 1.5995 | 0.9121 | | 0.676 | | | | |
| Streptococcus_sp_LPB0220 | 0.0220 | 0.0213 | 1.3247 | 0.22 | 0.694 | | | | |
| Bacteroides_unif | 0.0239 | 4.118 | 3.4275 | | 0.684 | | | | |

| | | | | | | | | | |
|---|---|---|---|---|---|---|---|---|---|
| ormis | | | | | | | | | |
| Paraprevotella_xylaniphila | 0.0249 | 1.1121 | 0.3984 | 0.9663 | 0.686 | | | | |
| Bacteroides_heparinolyticus | 0.0343 | 0.7408 | 0.2802 | | 0.672 | | | | |
| Collinsella_aerofaciens | 0.0449 | 3.6639 | 4.3819 | | 0.67 | | | | |
| Streptococcus_equinus | 0.0044 | -0.9569 | 0.5041 | 0.6782 | 0.741 | 0.610 to 0.873 | 0.0003 | 81.25 | 61.7 |
| Streptococcus_anginosus | 0.0079 | -1.1585 | 0.4881 | | 0.729 | 0.582 to 0.876 | 0.0022 | 56.25 | 85.11 |
| Streptococcus_gordonii | 0.0087 | -0.8940 | 0.0151 | | 0.718 | 0.583 to 0.853 | 0.0015 | 81.25 | 61.7 |
| Streptococcus_sp_FDAARGOS_192_12021 | 0.0091 | -1.0133 | 0.2898 | | 0.724 | 0.573 to 0.875 | 0.0037 | 87.5 | 48.94 |
| Streptococcus_sp_HSISS3 | 0.0259 | -1.1928 | 0.058 | | 0.693 | | | | |
| Clostridium_botulinum | 0.0305 | 0.2224 | -0.051 | | 0.686 | | | | |

**STable 9. Results of Uni- and Multivariate Cox regression for key phyla and genera**. Green color represents significant (<0.05) p-values and dark green color shows only a trend. Cox (M) analysis was performed only for taxa with a significant result (p<0.1) in Cox (U) analysis. For Lasso regression, Lasso coeff (n) is only shown for significant (p<0.05) predictors

| Taxa | Cox (U) p- | Cox (U) | Cox (U) | Cox (M) p- | Cox (M) | Cox (M) | Lasso |
|---|---|---|---|---|---|---|---|

| | value | Wald | Exp (b) | value | Wald | Exp (b) | coeff (n) |
|---|---|---|---|---|---|---|---|
| **Actinobacteria_Phylum** | 0.0928 | 2.825 | 1.3776 | >0.1 | | | 0.5961 |
| **Firmicutes** | 0.3056 | 1.0496 | 1.4132 | **0.0268** | 4.9034 | 2.118 | 0.2233 |
| **Barnesiella** | 0.2421 | 1.3683 | 0.7805 | >0.1 | | | 1.8581 |
| **Butyricimonas** | 0.0753 | 3.1644 | 0.734 | >0.1 | | | 1.0016 |
| **Bifidobacterium** | 0.0936 | 2.8116 | 1.2163 | >0.1 | | | ns |
| **Streptococcus** | **0.0082** | 6.9975 | 1.4995 | **0.0017** | 9.8447 | 1.7175 | ns |
| **Alistipes** | 0.1 | | | >0.1 | | | ns |
| **Methanosphaera** | 0.2473 | 1.3382 | 1.1274 | >0.1 | | | 1.2317 |
| **Buchnera** | **0.0314** | 4.6282 | 0.3189 | >0.1 | | | ns |
